# The Magnitude, Distribution Patterns, and Risk Factors of AIDS-Related Illnesses in Adults on Combined Antiretroviral Therapy in Malawi

**DOI:** 10.1101/2023.11.14.23298518

**Authors:** Hemson Hendrix Salema

## Abstract

This study describes the burden and spectrum of AIDS-related opportunistic diseases (ODs) in adults living with HIV (ALHIV) receiving combined antiretroviral therapy since the scale-up of free HIV treatment in Malawi. Data on AIDS-related ODs were abstracted from the 2004-2015 medical records and analysed using logistic regression models. 9,953 patients, with median age of 40 years (interquartile range (IQR): 33–48 years), of which 60.8% were females and 86.8% were within active sexual age (15-54 years), were included in the analysis. 65.1% were urban residents and 59.6% were from Southern region. 12,814 AIDS-related opportunistic episodes were extracted: 56.5% were prevalent AIDS-defining conditions and 43.5% were incident and recurrent cases. 7.7% of patients did not manifest any OD. Mycobacterial (36.3%), bacterial (20.8%), fungal (15.8%) and viral (15.1%) aetiological agents were most identified common pathogens. Tuberculosis (34.4%), bacterial pneumonia (11.2%), nontyphoid Salmonella bacteraemia (9.6%), HIV-wasting syndrome (8.9%), candidiasis (6.8%), isosporiasis (5.2%), pneumocystis pneumonia (4.2%) and cryptococcal meningitis (3.7%) were overall, most prevalent ODs. In the multivariate regression, healthcare facility types, gender, age-group, residential area, CD4 count, viral load, treatment initiation eligibility criteria and socio-economic status were identifiable risk factors of AIDS-related opportunistic diseases. The high prevalence of ODs among ALHIV in Malawi regardless of ART status requires establishing and strengthening of efficient strategies for effective prevention, early detection, and proactive management of AIDS-related ODs across all patient groups, regions, and health facilities.

**What this study adds**
This study provides insights into the scale and magnitude, the profile, the epidemiological and geographical distribution patterns, as well as associated risk factors of AIDS-related opportunistic diseases among adults living with HIV in Malawi since the introduction and scale-up of free antiretroviral therapy.

## BACKGROUND

The human immunodeficiency virus (HIV) infection remains a major global health challenge. Thus; since discovered, 74.9 million people have been affected, 37.9 million are living with HIV and 32.8 million have died of acquired immunodeficiency syndrome (AIDS) [1, 2]. HIV causes progressive depletion of CD4 T lymphocyte cells leading to debilitating, life-threatening opportunistic diseases (ODs) [3-8]. According to earlier epidemiological studies, over 90% of ODs are responsible for AIDS-related deaths [9] which have direct link to pathogenic virulence, degree of host immunity and antimicrobial prophylaxis usage [4]. ODs are related to reduced quality of life, increased stigma, and high medical costs [4, 10] and act as precursors to poverty and major causes of morbidity and mortality even post-ART initiation [11, 12] besides other attributed factors [13-19].

Although HIV pathophysiological pattern is similar in most patients, the patterns of ODs tend to differ across regions [20-22]. Hence, disparities in OD magnitude and profile between high-income and low-income and within the low-middle income countries exist [23-25]. Serious ODs which rarely manifest among PLHIV from developed nations are major morbidity and mortality causes within resource-limited nations [5, 16, 26-29].

Antiretroviral therapy (ART) uptake in Malawi has been slow compared to expert estimates of the country’s ART needs. Between 2003 and 2005, national ART uptake was an estimated 4,000, 13,000 and 28,110 only respectively, against the World Health Organisation (WHO) estimated 130000, 140000, and 169000 [30-32]. However, since the scale-up of free ART in 2004, Malawi has had copious achievements in HIV service expansion. ART clinics grew from 23 in 2004 to 716 in 2015 [30-32] and by December 2015, 68% (N=872,567) of approximately 1.1 million Malawi’s HIV population were receiving ART [33-35] thereby averting roughly 275,000 deaths and gaining 1.4 million life-years of mostly young adults in their peak productive life [36]. Moreover, indicators progressively show that Malawi is on course with the WHO’s 90-90-90 targets [35, 37, 38] such that by Dec 2018, 90% of PLHIV knew their status, of which 87% were on treatment and 89% were virally suppressed [39, 40]. However, despite tremendous gains, the incidence and prevalence of AIDS-related ODs in Malawi remain high irrespective of ART status [32, 41-48]. This study explores the scale, profile, and distribution pattern of AIDS-related ODs in Malawi since ART scale-up.

## METHODS

### Study setting, design, and participants

This study was conducted in eight randomly selected healthcare facilities across Malawi. Electronic patient’s medical records called ART Master cards, were used as sampling frames. A retrospective chart review was conducted between March and May 2016. Participants were all persons living with HIV aged 15+ years that commenced treatment using standardised protocols between Jan 2004 and Dec 2015. Study outcomes included all AIDS-defining and recurrent opportunistic diseases.

Study times were structured into two time-periods corresponding to major milestones in HIV management relating to treatment initiation eligibility criteria. The first period was designated “early-ART 2004-2011”. This referred to period between 2004-2011 when ART was initiated to only severely ill patients with either clinical stage III or IV of HIV disease or, nadir CD4 count <200 cells/μL [32, 49-53]. The second period; “late-ART 2011-2015”, referred to period between 2011-2015 when ART access was expanded to include patients with CD4 count ≤350 cells/μL regardless of WHO clinical stage [49-52].

### Sample size and sampling procedure

The sample size was determined from the following formula recommended for prevalence studies [54-59]:

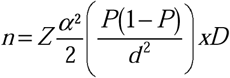

The estimated sample size of (n=8,654) was adjusted by 15% allowance to compensate for poor documentation resulting in final sample size of (n=9,953). A three-stage sampling method was used to randomise health facilities and medical records. Health facilities were selected from the list of 716 existing ART facilities as recorded in the Ministry of Health HIV Unit 2015 report [32, 60]. Participating sites were sampled using probability proportional to size from primary sampling units from the simple formula…. *K=N/n* [61, 62]. At each ART centre, exclusion criteria were applied to the primary data frame [(*N* _1_)], to form secondary sampling frame, [*N* _2_]; a stratum of eligible patients. The sampling procedure was replicated to randomly sample potential participants from *N* _2_. Anonymised patients’ serial numbers [from (*N* _2_)] were listed from which… *n*_1_, *n* _2_, *n* _3_, *n* _4_, *n* _5_, *n*_*x*_ serial numbers were randomly selected.

### Data collection and analysis procedures

Data were collected using an electronic tool designed to capture data on both prevalent and incident AIDS-associated clinical conditions, associated risk factors and non-identifiable demographic details.

Socio-demographic and clinical parameters were analysed descriptively. Frequency and proportions were expressed as absolute numbers and percentages. Statistically significant differences between patients with and without specific OD were compared using Chi-square test. The mean (with standard deviation, ±SD) and median (with interquartile range, IQR) between the two patient groups were compared using two-sample *t*-test and one-way analysis of variance. Logistic regression analyses were used to determine associations between variables and OD manifestations, and findings were reported as odds ratios with 95% confidence intervals (OR 95% CI). Factors with *p* ≤ 0.25 from the crude analysis were entered into final model. Significance level was set by two-tailed alpha (α) of 0.05. The multicollinearity tolerance test, Hosmer-Lemeshow test, and the linktest were used to determine goodness-of-fit of final models. All statistical procedures were performed using Stata special edition for windows v15.1.

## RESULTS

### Participants socio-demographic characteristics

Sample disposition details are presented in Table 1. A total of 9,953 medical records of treatment-exposed patients that initiated ART between Jan 2004 and Dec 2015 were abstracted – achieving a 100% planned retrieval rate. Participants mean (±SD) age was 40.7±12.2 years. Most were females and relatively younger than males; mean age: 38.8 vs 43.6 years respectively (*p<*0.0001), and 86.8% were within active sexual age (15-54 years). Whereas most patients were urban residents, majority were recruited from Southern region (59.6%) compared to Northern and Central regions. 78.8% were married and 13.9% were either divorced or widowed and only 7.4% were ever-single. Majority (67.7%) were recruited from tertiary-level healthcare facilities, 7.2% had no education, and 55.3% were employed.

**Table 1:** Sample Disposition: PLHIV on Combined ART; Malawi, 2004-2015.

| Characteristic | Number (%) | Characteristic | Number (%) |
| --- | --- | --- | --- |
| **Care type** |  | **Marital status** |  |
| Primary care | 735 (7.38) | Previously married | 1,380 (13.87) |
| Secondary care | 2,482 (24.94) | Married (partnership) | 7,837 (78.74) |
| Tertiary care | 6,736 (67.68) | Single (never married) | 733 (7.36) |
| Total | 9953 (100.00) | Total | 9953 (100.00) |
| **Gender** |  | **Education levels** |  |
| Female | 6,055 (60.84) | No education | 713 (7.16) |
| Male | 3,898 (39.16) | Primary | 3,349 (33.65) |
| Total | 9953 (100.00) | Secondary | 3,260 (32.75) |
| **Age-range (years)** |  | Higher | 2,631 (26.43) |
| 15-19 (Adolescents) | 358 (3.60) | Total | 9953 (100.00) |
| 20-39 (Young adults) | 4,501 (45.22) | **Employment status** |  |
| 40-54 (Mid-Aged Adults) | 3,785 (38.03) | Employed | 5,506 (55.32) |
| 55+ (Older Adults) | 1,306 (13.12) | Self-employed | 4,447 (44.68) |
| Total | 9953 (100.00) | Total | 9953 (100.00) |
| **Residential area** |  | **Occupation classification** |  |
| Rural | 2,047 (20.57) | Manual jobs | 1,658 (16.66) |
| semi-urban | 1,427 (14.34) | Others job types | 4,949 (49.72) |
| Urban | 6,477 (65.08) | White collar jobs | 3,346 (33.62) |
| Total | 9953 (100.00) | Total | 9953 (100.00) |
| **Regional area** |  | **ART-initiation criteria** |  |
| Central Region | 2,441 (24.53) | CD4+ count | 2,475 (24.87) |
| Northern Region | 1,585 (15.92) | Option-B+/PMTCT | 261 (2.62) |
| Southern Region | 5,927 (59.55) | WHO III | 4,781 (48.04) |
| Total | 9953 (100.00) | WHO IV | 2,436 (24.48) |
| **Guardian escort** |  | Total | 9953 (100.00) |
| None (no guardian) | 1,471 (14.78) |  |  |
| Yes (with guardian) | 8,481 (85.21) |  |  |
| Total | 9953 (100.00) |  |  |

### Baseline clinical characteristics of study participants

Table 2 summarises participants’ clinical characteristics. All patients were once commenced on ART under standardised treatment protocols. 72.5% commenced treatment with advanced HIV disease: WHO clinical stage III and IV and 27.5% were asymptomatic – of which, 24.9% had low CD4 count below recommended threshold and 2.6% were either breastfeeding or pregnant; the Option-B+/PMTCT criterion. Overall, 67.1% of patients were commenced with Efavirenz-based regimen compared to Nevirapine-based or non-standard ART regimens. 95.4% were continuing with first-line regimens while 3.3% had been switched to second-line regimens; the boosted-Ritonavir regimens – meaning they had proven first-line ART failure.

**Table 2:** Clinical Characteristics of Adults living with HIV on ART; Malawi, 2004-2015.

| Characteristic | Number (%) | Characteristic | Number (%) |
| --- | --- | --- | --- |
| **ART-initiation criteria** |  | **Maintenance ART regimen** |  |
| CD4+ count | 2,475 (24.87) | Nevirapine-based | 2,416 (24.27) |
| Option-B+/PMTCT | 261 (2.62) | Efavirenz-based | 7,080 (71.13) |
| WHO III | 4,781 (48.04) | Boosted-Ritonavir | 327 (3.29) |
| WHO IV | 2,436 (24.48) | Non-std. | 130 (1.31) |
| **CD4 initial results** |  | **Contraception (Barrier)** |  |
| 0-250 cells/μL | 6,956 (69.89) | Yes | 7,087 (71.20) |
| 250+ cells/μL | 2,988 (30.02) | No | 2,866 (28.80) |
| **CD4 recent results** |  | **HIV duration** |  |
| 100-500 cells/μL | 3,125 (31.40) | 1-2 yrs. | 1,342 (13.48) |
| 501-1,000 cells/μL | 5,201 (52.26) | 3-5 yrs. | 2,610 (26.22) |
| 1000+ cells/μL | 1,625 (16.33) | 6-10 yrs. | 4,440 (44.61) |
| **Initial VL test results** |  | 10+ yrs. | 1,561 (15.68) |
| <3.0 Log<sub>10</sub> copies/mL | 798 (8.02) | **ART duration** |  |
| >3.0 Log<sub>10</sub> copies/mL | 9,047 (90.89) | 1-2 yrs. | 2,154 (21.64) |
| **Follow-up VL tests results** |  | 3-5 yrs. | 3,243 (32.58) |
| <1.6 Log<sub>10</sub> copies/mL | 3,992 (40.11) | 6-10 yrs. | 3,933 (39.52) |
| 1.6-3.0 Log<sub>10</sub> copies/mL | 3,048 (30.62) | 10+ yrs. | 623 (6.26) |
| >3.0 Log<sub>10</sub> copies/mL | 2,913 (29.27) | **Clinical latency range** |  |
| **Initial ART regimen** |  | 1-2 years | 6,964 (69.97) |
| Nevirapine-based | 2,840 (28.53) | 3-5 years | 2,259 (22.70) |
| Efavirenz-based | 6,678 (67.10) | 6+ years | 730 (7.33) |
| Non-std. | 435 (4.37) | **Patient-outcome** |  |
|  |  | Treatment adherent | 6,758 (67.90) |
|  |  | Defaulters & LTFU | 2,018 (20.28) |
|  |  | Died | 1,177 (11.83) |

CD4 cell count, and viral load test data were extracted at two time-points: as initial baseline and as follow-up results (the latter were the most recent available test results). Overall, the mean (±SD) initial baseline CD4 count was 145.1±95.1 cells/μL. Most patients had severe immunosuppression; with initial baseline CD4 count <250 cells/μL. Following treatment, CD4 count results improved considerably in most patients to normal healthy adult levels; overall mean (±SD) follow-up CD4 count was 648±238.6 cells/μL, and in 68.6% of patients, the follow-up CD4 counts were >500 cells/μL. Similarly, most patients had severe viral failure when commenced treatment. The mean (±SD) initial baseline viral load (VL) was 6.0±4.3 Log_10_ copies/mL, and majority had >4.9 Log_10_ copies/mL. Likewise, the mean follow-up VL was relatively lower following treatment; 5.1±3.8 Log_10_ copies/mL and up to 70.7% were virally suppressed (VL<3.0 Log_10_ copies/mL) of which, (N=3,992) had undetectable VL results of <1.6 Log_10_ copies/mL.

### The magnitude and profile of AIDS-associated opportunistic conditions

A cumulative total of 12,814 episodes of AIDS events (opportunistic diseases) were recorded from the (N=9,953) enrolled patients. 58.9% (N=7,559) were prevalent AIDS-defining clinical conditions (ADCCs) that manifested in treatment-naïve patients and 41.1% (N=5,255) were incident AIDS-recurrent conditions in treatment-experienced patients. The overall period prevalence of prevalent ADCC between 2004 and 2015 was an estimated 72.6% in contrast to an estimated 55.8% for treatment-experienced patients with repeated occurrences of some conditions. Of the six aetiological agents responsible for causing the opportunistic episodes, mycobacterial were most prevalent (36.3%) while neoplastic conditions were the least (Fig. 1). In 7.7% (N=761) of patients, no AIDS event had occurred, hence, remained asymptomatic. Further analyses showed that 67.9% were treatment compliant, 20.3% were non-compliant; (i.e., either defaulters, stopped treatment, or lost to follow-up), and 11.8% had died.

**Figure 1:**
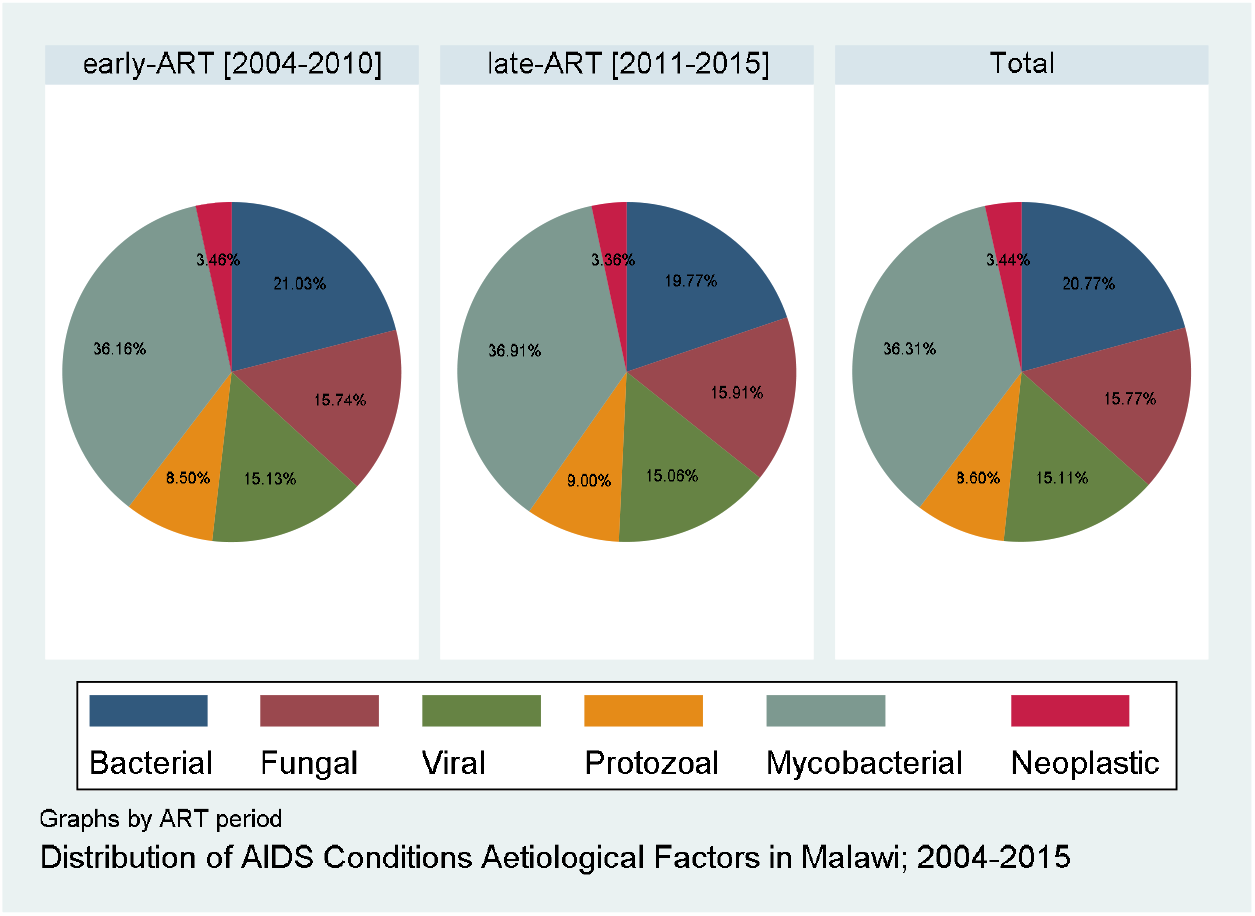
Distribution of aetiological factors responsible for causing AIDS-related opportunistic diseases during the study’s early-ART period (2004-2010) and late-ART period (2011-2015) in Malawi.

The frequency of individual OD occurring ranged from 1 t**o** 4 times per patient. 60.3% had (only) single, 29.2% had dual, 2.1% and 0.9% had triple and quadruple OD occurrence respectively (Fig. 2) whereas 7.6% had never manifested any OD.

**Figure 2:**
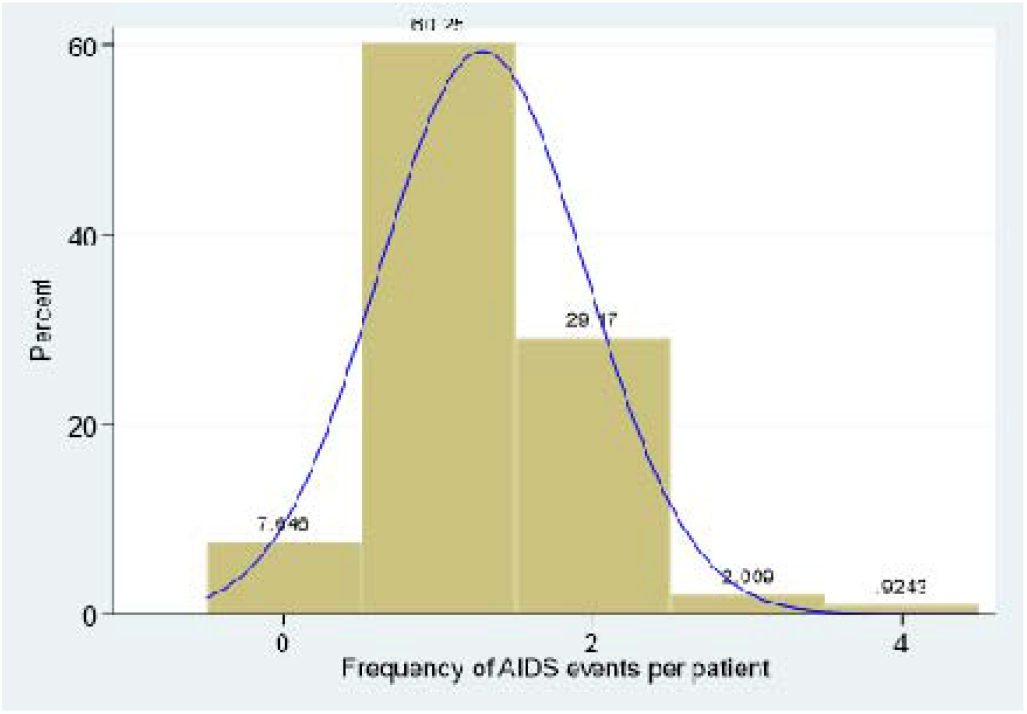
Frequency of AIDS opportunistic disease episodes per patient.

Most ODs were diagnosed in tertiary health facilities (67.7%) while primary facilities accounted for only 7.4%. Figure 3 illustrates yearly frequencies of new HIV cases and against the number of OD episodes stratified by year of admission into HIV care. 16.4% (N=1,636) of HIV cases were diagnosed before 2004 which was outside the study period. Within the study period, more HIV cases were registered into care between Jan 2005 and Dec 2007 and most patients commenced treatment in 2006 (12.9%, N=1,286) but were least in 2015 (1.6%).

**Figure 3:**
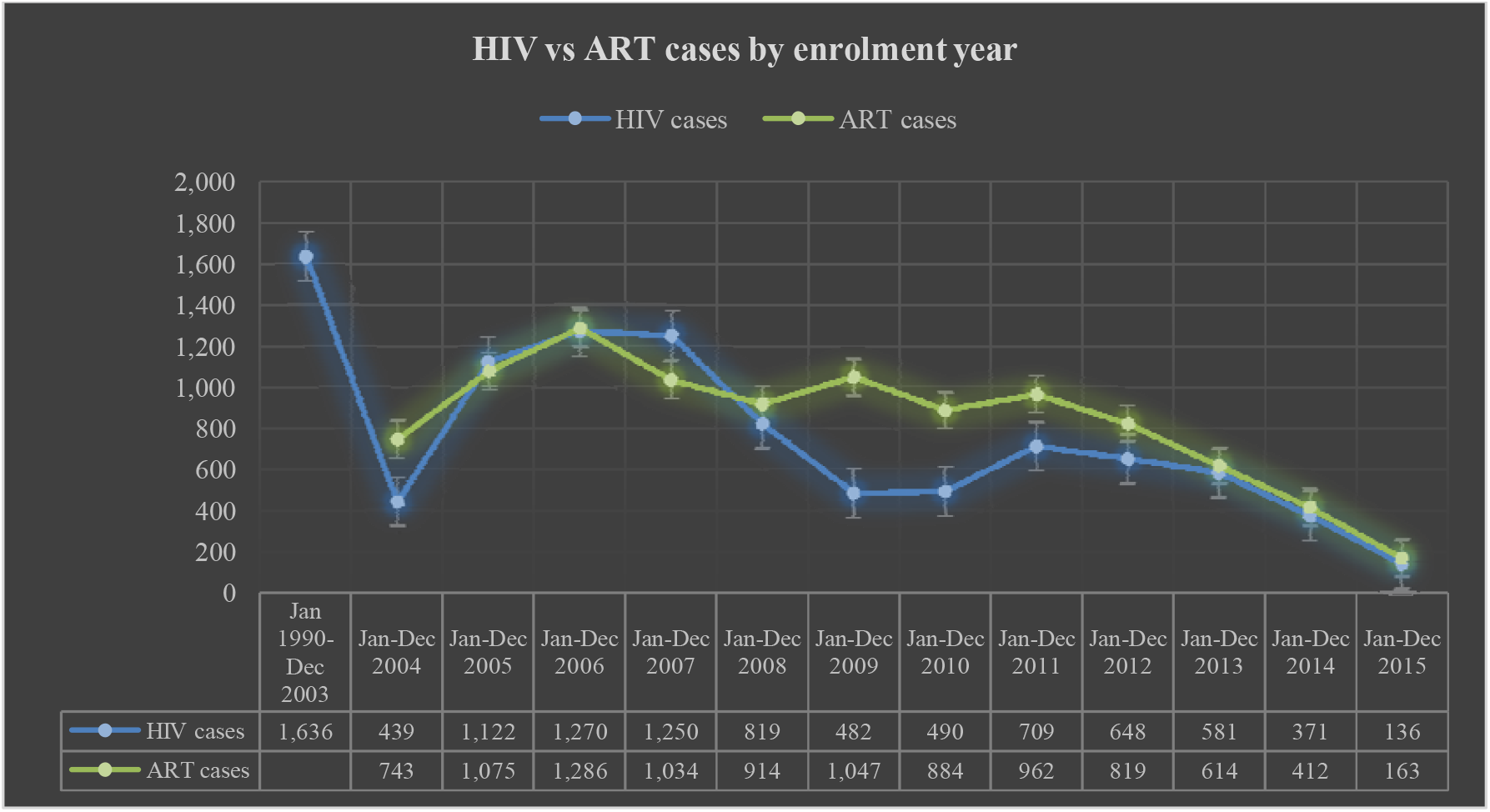
Frequency of yearly new HIV cases and compared with frequency of opportunistic disease episodes stratified by enrolment year into care from 2004 to 2015 (including HIV cases enrolled into care between 1990 and 2003).

### The prevalence of opportunistic diseases

Table 3 shows the frequency of ODs stratified by ART periods and healthcare facility types and the combined probability of each OD diagnosis in primary and secondary facilities against tertiary health facilities. Overall, tuberculosis (TB) was most prevalent OD (35.4%) of which the pulmonary type (PTB) was more prevalent (20.4%) than extrapulmonary (EPTB) type (13.9%). Other highly prevalent conditions included bacterial pneumonia (11.5%), non-typhoid salmonella bacteraemia (9.9%), HIV wasting syndrome (9.1%), candidiasis (7.1%), isosporiasis (5.2%), pneumocystis pneumonia (4.1%) and cryptococcal meningitis (3.7%).

**Table 3:** Frequency of ODs Stratified by ART-Initiation Period and Health Facility Type.

|  | Overall | Early-ART<br>(2004-2011) | Late-ART<br>(2011-2015) | Tertiary<br>Healthcare<br>Facilities | Secondary<br>Healthcare<br>Facilities | Primary<br>Healthcare<br>Facilities | OR (95% CI):<br>Primary &<br>Secondary Rates<br>Vs<br>Tertiary<br>H/Facilities |
| --- | --- | --- | --- | --- | --- | --- | --- |
| AIDS condition | Frequency<br>(%) | Frequency<br>(%) | Frequency<br>(%) | Frequency<br>(%) | Frequency<br>(%) | Frequency<br>(%) |  |
| M. Tuberculosis (overall) | **4392 (34.39)** | **3482 (34.20)** | **910 (35.14)** | **3027 (25.75)** | **1019 (24.53)** | **346 (27.20)** | **1.11 (1.02, 1.21)\*\*** |
| Pulmonary tuberculosis | 2611 (20.44) | 2067 (20.30) | 544 (21.00) | 1853 (21.23) | 553 (17.63) | 205 (22.14) | 1.23 (1.12, 1.36)** |
| EPTB | 1781 (13.95) | 1415 (13.89) | 366 (11.13) | 1174 (13.45) | 466 (14.86) | 141 (15.23) | 0.91 (0.81, 0.98)** |
| Bacterial pneumonia | 1431 (11.53) | 1156 (11.35) | 275 (10.62) | 937 (7.97) | 406 (9.77) | 88 (6.92) | 0.89 (0.79, 0.98)** |
| NTS bacteraemia | 1222 (9.85) | 985 (9.67) | 237 (9.15) | 793 (6.75) | 352 (8.47) | 77 (6.05) | 0.87 (0.76, 0.98)** |
| HIV-wasting syndrome | 1140 (9.02) | 914 (8.98) | 226 (8.73) | 738 (6.28) | 314 (7.56) | 88 (6.92) | 0.86 (0.76, 0.98)** |
| Candidiasis | 870 (7.01) | 694 (6.82) | 176 (6.80) | 588 (5.00) | 214 (5.15) | 68 (5.35) | 0.99 (0.85, 1.15) |
| Isosporiasis | 669 (5.17) | 527 (5.18) | 142 (5.48) | 460 (3.91) | 167 (4.02) | 42 (3.30) | 1.05 (0.89, 1.24) |
| Pneumocystis pneumonia | 513 (4.02) | 407 (4.00) | 106 (4.09) | 364 (3.10) | 117 (2.82) | 32 (2.52) | 1.18 (1.09, 1.43)** |
| Cryptococcal meningitis | 475 (3.72) | 385 (3.78) | 90 (3.47) | 332 (2.82) | 114 (2.74) | 29 (2.28) | 1.11 (1.09, 1.36)** |
| Cryptosporidiosis | 376 (2.94) | 293 (2.88) | 83 (3.20) | 266 (2.26) | 81 (1.95) | 29 (2.28) | 1.61 (0.93, 1.45) |
| Herpes Simplex virus | 300 (2.35) | 247 (2.43) | 53 (2.05) | 198 (1.68) | 81 (1.95) | 21 (1.65) | 0.92 (0.73, 1.18) |
| Kaposi's Sarcoma (KS) | 298 (2.33) | 242 (2.38) | 56 (2.16) | 199 (1.69) | 72 (1.73) | 27 (2.12) | 0.96 (0.75, 1.22) |
| NTM | 245 (1.92) | 199 (1.95) | 46 (1.78) | 169 (1.44) | 56 (1.35) | 20 (1.57) | 1.06 (0.81, 1.40) |
| Cytomegalovirus infection | 160 (1.25) | 114 (1.12) | 46 (1.78) | 146 (1.24) | 10 (0.24) | 21 (1.65) | 5.07 (2.98, 8.79)** |
| Mycosis | 156 (1.22) | 116 (1.14) | 40 (1.54) | 138 (1.17) | 12 (0.29) | 6 (0.47) | 3.72 (2.27, 6.09)** |
| HIV-encephalopathy | 137 (1.07) | 108 (1.06) | 29 (1.12) | 100 (0.85) | 34 (0.82) | 3 (0.24) | 1.30 (1.18, 1.89)** |
| Invasive Cervical Cancer | 108 (0.85) | 82 (0.81) | 26 (1.00) | 81 (0.69) | 20 (0.48) | 7 (0.55) | 1.43 (1.39, 2.23)** |
| HIV-cardiomyopathy | 82 (0.64) | 67 (0.66) | 15 (0.58) | 52 (0.44) | 28 (0.67) | 2 (0.16) | 0.83 (0.53, 1.30) |
| HIV-nephropathy | 69 (0.54) | 54 (0.53) | 15 (0.58) | 49 (0.42) | 15 (0.36) | 5 (0.39) | 1.17 (0.70, 1.97) |
| Toxoplasmosis | 53 (0.42) | 45 (0.44) | 8 (0.31) | 40 (0.34) | 6 (0.14) | 7 (0.55) | 1.47 (1.38, 2.76)** |
| PML | 42 (0.33) | 36 (0.35) | 6 (0.23) | 29 (0.25) | 7 (0.17) | 6 (0.47) | 1.07 (0.55, 2.05) |
| Non-Hodgkin's lymphoma | 33 (0.26) | 28 (0.28) | 5 (0.19) | 21 (0.18) | 10 (0.24) | 2 (0.16) | 0.83 (0.41, 1.70) |
| **Totals** | **12,814**<br>**(100%)** | **10,181**<br>**(100%)** | **2,590**<br>**(100%)** | **8,727**<br>**(100%)** | **3,135**<br>**(100%)** | **926**<br>**(100%)** | **--** |

\*\* statistically significant odds ratio of primary & secondary health facilities' rates Vs tertiary health facilities
The table (above) shows rates of individual opportunistic diseases: overall rates, rates stratified by early- (2004-2011) and late- (2011-2015) ART periods and by existing healthcare facility types. Combined rates for primary and secondary health facilities are/were compared with those of tertiary facilities in an unadjusted regression. Statistically significant findings means the condition was more (or less) likely to manifest in patients attending tertiary than those attending primary / secondary healthcare facilities.
**Key:** EPTB: Extrapulmonary Tuberculosis; NTS: Nontyphoid Salmonella bacteraemia; NTM: Nontuberculous mycobacteria; PML: Progressive multifocal leukoencephalopathy

The estimated OD prevalence during both ART periods were mostly proportionally indifferent. For example, TB prevalence during early-ART was 34.2% compared to 35.1% in late-ART while candidiasis was 6.8% during both ART periods. Overall, most AIDS conditions tended to be diagnosed in tertiary than primary and secondary healthcare facilities combined. Thus, TB was 11% [OR=1.11, 95% CI: (1.02, 1.21)], cytomegalovirus infection was 5 times [OR=5.07, 95% CI: (2.98, 8.79)], mycosis was 3 times [OR=3.72, 95% CI: (2.27, 6.09)], toxoplasmosis was 47% [OR=1.47, 95% CI: (1.38, 2.76)], and HIV encephalopathy and ICC were 30% and 43% more likely to be diagnosed in tertiary healthcare facilities than primary/secondary tier combined.

## RISK FACTORS OF AIDS-ASSOCIATED OPPORTUNISTIC DISEASES BASED ON AETIOLOGICAL FACTORS

### Bacterial opportunistic diseases

Risk factors associated with bacterial ODs are given in Appendix A, Table 4. Bacterial pneumonia and nontyphoid Salmonella bacteraemia are the two recognisable bacterial ADCC. In the multivariate analysis, the risk of bacterial OD manifestation was increased by 34% in secondary compared to primary level facilities [aOR 1.34, 95% CI: (1.08, 1.66)], by 15% in urban compared to rural areas [aOR 1.15, 95% CI: (1.01, 1.30)], by 9% in Southern compared to central region [aOR 1.09, 95% CI: (1.07, 1.23)], and by two-folds and 98% in advanced HIV disease clinical stage III and stage IV respectively [aOR 2.22, 95% CI: (1.93, 2.55)], and [aOR 1.98, 95% CI: (1.71, 2.31)], compared to low CD4 count. The risk was likewise increased by 92% with high initial baseline VL of >3.0 Log_10_ copies/mL compared to VL <3.0 Log_10_ copies/mL and by 24% with defaulting treatment compared to continuing with ART (aOR 1.24, 95% CI: (1.02, 1.49). Comparatively, the probability of bacterial OD occurrence was decreased by 21% in Northern region [aOR 0.79, 95% CI: (0.67, 0.94)], and by 12% among self-employed patients [aOR 0.88, 95% CI: (0.79, 0.97)], by 10% with follow-up CD4 count results of >1000 cells/μL compared to CD4 count <500 cells/μL, [aOR 0.90, 95% CI: (0.76, 0.96)].

### Fungal opportunistic diseases

Candidiasis, pneumocystis pneumonia, cryptococcal meningitis and mycosis, are the four fungal ADCCs. In the multivariate analysis (Appendix A, Table 4), the probability of AIDS-related fungal ODs were increased by 33% in secondary [aOR 1.33, 95% CI: (1.26, 4.91), and by over four-folds in tertiary healthcare facilities [aOR, 4.42, 95% CI: (1.38, 14.07)], compared to primary facilities. The risk was likewise increased by 15% and 52% in urban and semi-urban areas, respectively [aOR 1.15, 95% CI: (1.09, 1.93) and aOR=1.52, 95% CI: (1.48, 2.76)] compared to rural areas, by 18% and 85% in HIV clinical stage III and stage IV respectively (during ART commencement) [aOR 1.18, 95% CI: (1.03, 1.36)], and [aOR 1.85, 95% CI: (1.59, 2.16)] compared to low CD4 count, by 70% with initial baseline VL >3.0 Log_10_ copies/mL compared to <3.0 Log_10_ copies/mL, [aOR 1.70, 95% CI: (1.23, 2.35)], and by 78% with defaulting treatment [aOR 1.78, 95% CI: (1.64, 1.97)]. The risk was nonetheless decreased by 49% with initial baseline CD4 count >250 cells/μL compared to <250 cells/μL, [aOR 0.59, 95% CI: (0.52, 0.66)], and by 32% and 39% with follow-up CD4 count between 500-1000 cells/μL and over 1000 cells/μL, respectively [aOR 0.61, 95% CI: (0.38, 0.98)] and [aOR 0.68, 95% CI: (0.38, 0.73)] compared to <500 cells/μL.

### Viral opportunistic diseases

Risk factors associated with AIDS-defining viral ODs are summarised in Appendix B, Table 5. The risk of viral ADCC was increased by 3 and 4 times in secondary and tertiary healthcare facilities [aOR 3.72, 95% CI: (1.88, 15.79)] and [aOR 4.72, 95% CI: (1.15, 19.32)], compared to primary facilities. Equally, the risk was increased by 23% and 13% in semi-urban and urban residents, [aOR 1.23, 95% CI: (1.02, 1.48) and aOR 1.13, 95% CI: (1.08, 2.48)] compared to rural residents, by 14% in Southern region, [aOR 1.14, 95% CI: (1.01, 1.30)] compared to Central region, by 43% and 75% in advanced HIV disease stage III and stage IV respectively, [aOR 1.43, 95% CI: (1.23, 1.65), and [aOR 1.75, 95% CI: (1.49, 2.06)] compared to low CD4 count at treatment initiation, and by 92% in initial baseline VL >3.0 Log_10_ copies/mL [aOR 1.92, 95% CI: (1.67, 2.28)] compared to VL <3.0 Log_10_ copies/mL. Moreover, the risk was increased by two-folds in severe follow-up VL >3.0 Log_10_ copies/mL [aOR=2.23, 95% CI: (1.82, 2.55)], compared to undetectable VL but was decreased by 35% in Option-B+/PMTCT treatment initiation criterion, [aOR 0.65, 95% CI: (0.42, 0.91)] compared to low CD4 count.

### Mycobacterial opportunistic diseases

Tuberculosis and nontuberculous mycobacterium are the only recognised mycobacterial ADCCs. The multivariate logistic analysis (Appendix B, Table 5), showed that Northern region was associated with 8% increased risk of AIDS-related mycobacterial ODs, [aOR 1.08, 95% CI: (1.04, 1.23)] compared to Central region. Equally, the risk was increased by 16% and 23% in primary and higher education levels respectively, compared to patients without education (*p<*0.05), by 3-folds and 73% in advanced HIV clinical stage III and IV at treatment initiation respectively, [aOR 3.13, 95% CI: (2.80, 3.49)], and [aOR 1.73, 95% CI: (1.1.53, 1.96)] compared to low CD4 count, by 21% in initial baseline VL >3.0 Log_10_ copies/mL compared to <3.0 Log_10_ copies/mL and by 51% with non-standard ART regimen [aOR 1.51, 95% CI: (1.03, 2.19)], compared to Nevirapine-based maintenance regimens. In contrast, the risk was reduced by 25% with initial baseline CD4 count >250 cells/μL compared to <250 cells/μL, [aOR 0.75, 95% CI: (0.55, 0.96)], by 23% and 15% with secondary and tertiary healthcare facilities, [aOR 0.77, 95% CI: (0.64, 0.92) and aOR 0.85, 95% CI: (0.72, 0.91)] compared to primary level, by 55% in Option-B+/PMTCT criterion [aOR 0.45, 95% CI: (0.32, 0.64)] compared to low CD4 count at treatment initiation.

### Protozoal opportunistic diseases

Cryptosporidiosis, isosporiasis and toxoplasmosis are the recognisable ADCCs. In the multivariate logistic analysis (Appendix C, Table 6), the risk of AIDS-defining protozoal ODs was increased by 5% in males [aOR 1.05, 95% CI: (1.02, 1.25)] compared to females, by 36% and 26% in advanced HIV disease clinical stage III and stage IV respectively [aOR 1.36, 95% CI: (1.1.13, 1.64)] and [aOR 1.26, 95% CI: (1.02, 1.56)] compared to low CD4 count at ART initiation, by 15% in initial baseline VL >3.0 Log_10_ copies/mL [aOR 1.15, 95% CI: (1.07, 1.79)] compared to VL <3.0 Log_10_ copies/mL, and by 78% in non-standard ART [aOR 1.78, 95% CI: (1.08, 2.93)] compared to nevirapine-based maintenance regimens. In contrast, the risk was statistically significantly decreased by 35%, 32% and 44% in 20-39 years, 40-54 years, and 55+ years age-group respectively compared 15-19 years, by 19% and 23% in follow-up CD4 count of between 500-1000 cells/μL and >1000 cells/μL compared to <500 cells/μL, and by 17% in Efavirenz-based [aOR 0.83, 95% CI: (0.71, 0.97)] compared to Nevirapine-based maintenance ART regimens.

### Malignant neoplastic opportunistic diseases

Malignant Neoplastic ODs include invasive cervical cancer, Kaposi’ sarcoma and non-Hodgkin lymphoma. In the multivariable analysis (Appendix C, Table 6), the risk of malignant neoplastic ODs was increased by 86% and 94% in secondary and tertiary healthcare facilities [aOR 1.86, 95% CI: (1.55, 2.35) and aOR 1.94, 95% CI: (1.63, 2.42)], compared to those primary health facilities, by 3-folds in advanced HIV disease; clinical stage IV [aOR 3.24, 95% CI: (2.46, 4.27)] compared to low CD4 count, and by 74% in initial baseline VL >3.0 Log_10_ copies/mL compared with <3.0 Log_10_ copies/mL, [aOR 1.74, 95% CI: (1.39, 2.40)]. Contrastingly, the risk of neoplastic ODs was statistically significantly decreased by 27% in males compared to females, [aOR 0.73, 95% CI: (0.56, 0.95)], and by 77% in Option-B+/PMTCT treatment initiation criterion [aOR 0.23, 95% CI: (0.06, 0.96) compared to low CD4 count.

## DISCUSSION

This study elucidates the scale, magnitude, distribution patterns, and risk factors of AIDS-related ODs among treatment-exposed adults living with HIV (ALHIV) in Malawi using the 2004-2015 nationally representative data. To my knowledge, this is the first study using such data to explore prevalence and associated factors of AIDS-associated ODs and the results indicate significant burden of ODs among PLHIV.

We abstracted both prevalent and incident conditions that are typically ADCC from enrolled patients during the 11-year data generation period. OD frequency ranged from 1 to 4 times per patient such that 60.3%, 29.2%, 2.1% and 0.9% of patients had single, dual, triple, and quadruple OD episodes.

The overall cumulative OD period prevalence in the study was remarkably high (128.3%) due to repeated occurrences. This finding compares with 120.9% overall cumulative prevalence from Taiwan University Hospital study which reported 1,263 OD episodes from (*n=*1,044) patients [17], but is much lower than 268.1% from The AIDS Support Organisation (TASO) HIV facilities in Uganda which reported 291,168 OD episodes from (*n=*108,619) HIV patients from 2001 to 2013 [63].

Of the (N=12,814) OD episodes identified in this study, 59.1% and 40.9% were prevalent and incident cases respectively. Therefore, the (period) prevalence of incident ODs among treatment-experienced adults between 2004 and 2015 was an estimated 55.8% with repeated occurrences. This finding is comparable to 48.1% reported in Hiwot Fana Hospital [7]; and 42.8% prevalence reported in Debre Markos Hospital [19] both from Ethiopia. The finding is likewise comparable to 47.6% prevalence reported from both Taiwan University Hospital [17] and from Tshepang Clinic South Africa [64]. However, it is relatively higher than 19.7% prevalence reported in Gondar Hospital [65], 22.4% prevalence reported in South East Nigeria [66] and 30% prevalence reported in Bamrasnaradura institute Thailand [67].

The present study summarised clinical data representing 1.0% of all PLHIV in Malawi and 1.3% of treatment-exposed HIV infected adults. Most were females (60.8%) and majority were urban residents (65.1%) within reproductive group of 15-49 years (78.2%). Moreover, 78.4% were married, 40.8% had low education, and 49.7% had low socio-economic status although the majority (55.3%) were employed. The nature of patients in the current study compares with others from sub-Saharan Africa (SSA) region including Uganda, Ethiopia and South Africa [7, 12, 19, 63, 66, 68, 69]. In a cross-sectional review from Uganda, 64% were women, 77% had low education and 76% had low socio-economic status [63]. Likewise in Nigeria, 65.8% were women and 50.4% had low socio-economic status [66] while a cross-sectional study from Ethiopia; 61.8% were women, 96.2% were of reproductive group, 50.8% were married and 68.6% had low education ([12]. In these studies, women had consistent higher proportions; implying that either, men probably are under-represented in most SSA region ART programmes (resulting into less favourable outcomes) or that women remain disproportionately affected by HIV/AIDS.

Of all OD episodes in the current study, opportunistic infections (OIs) accounted for 96.9% compared to 3.1% of malignant neoplastic opportunistic conditions. This finding is consistent with that from Uganda [70] and signifies that OIs; other than malignant opportunistic neoplasms, are principal causes of morbidity and mortality among ALHIV in SSA. Tuberculosis prevalence in the current study was comparatively high (35.4%) compared to findings from other studies elsewhere [7, 12, 19, 63, 66, 68, 69]. This observed difference was due to high TB rates in Malawi which had worsened with the advent of HIV [48, 71-75]. TB remains most prevalent in Malawi with PTB affecting roughly 11,000 people annually [42]. Besides, a considerable proportion of TB suspects are AAFB ‘smear negative’ [76].

One familiar condition called atypical disseminated leishmaniasis, was not seen in this study. While this may suggest (the) absent of Leishmania parasites in Malawi, lack of diagnostic capability may be the principal reason. To date, only two cases; both diagnosed at post-mortem and with travel history to Tanzania, have been recorded with the first reported in 1979 [77].

Expectedly, the burden of ODs during early-ART era (2004-2011), was indeed high; (79.8%) compared to 20.2% in late-ART era (2011-2015). During both periods, mycobacterial agents were common aetiological factors, followed by bacterial, fungal, and viral agents (Fig 3). Most prevalent conditions during early-ART era were TB (34.2%), bacterial pneumonia (11.4%), nontyphoid Salmonella (9.7%), HIV wasting syndrome (8.9%), candidiasis (6.8%) and chronic isosporiasis (5.2%). This pattern was similar with that of late-ART era and compared with findings from other studies across SSA region [7, 19, 65, 66].

### Study limitations & strengths

To my knowledge, this is the first study to collate and aggregate national representative data on the AIDS burden in treatment-exposed PLHIV in Malawi. The study used sufficient data with wider geographical coverage and involved all existing healthcare facility levels. This study has shown the estimated national prevalence of different AIDS-related conditions affecting adult HIV patients, revealed their types, magnitudes, distribution patterns and factors associated with their occurrences – in a way to inform policy and practice on managing ODs. Besides, the study has attempted to present the true picture of common ODs prevalent among HIV positive adults in Malawi by including treatment-naïve patients eligible for ART regardless of prophylaxis status. Each patient was then followed-up for as long as they remained in care – hence, able to estimate both prevalent and incident OD prevalence.

However, the study used pre-existing medical records hence, subject to limitations which may emanate from incomplete recording of vital clinical information and diagnostic capability. Thus, the study used data collected not primarily for medical research, hence, some variables potentially vital and complementary in interpretation of results like ART adherence, prophylaxis treatment received, functional status and clinical surrogate markers, were either completely absent or missing in some patients. It is therefore imperative that future cohort studies should target such data. Additionally, most healthcare facilities had no capability and competence to readily perform specimen cultures or confirmatory diagnosis of ODs such that certain conditions were clinically diagnosed thereby potentially impacting diagnosis accuracy. Thirdly, the study did not assess and ascertain pre-ART burden of ODs prior to the 2004 nationwide rollout of free ART. Hence, variations and expected declines in OD burden resulting from ART could not be ascertained. Fourthly, certain ODs were not captured in some healthcare facilities probably due to inadequate diagnostic capability. Besides, a small proportion of TB cases (N=109, 2.5%) were clinically unclassified when abstracted, hence, analysed as PTB for consistency. Finally, some suspected ODs seen in lower health facilities (primary and secondary) and referred to tertiary health facilities due to limited resources were often underestimated or missed as not all cases reached the referral centres.

## CONCLUSION

This study has demonstrated the pervasiveness of prevalent and incident opportunistic diseases among ALHIV in Malawi and shown that they remain a reckoning clinical and public health challenge. The magnitudes and profiles of ODs have shown to vary by different factors such as type of ODs, ART-era, age-group, gender, and geographical areas. Mycobacterial pathogens remain dominant primary causes of serious ODs in Malawi. All ODs identified in the study warrant particular attention – not just the most prevalent ones at the expense of conditions with relatively low prevalence. Additionally, independent predictors strongly associated with serious ODs merit special consideration especially when screening for disease HIV progression. Screening and prevention strategies for HIV and serious ODs require redesigning, monitoring, and prioritising. The need to strengthen strategies and care models that highlight early identification of new HIV cases and the need for nationwide accelerated implementation of test-and-treat approach cannot be overemphasised. Prophylaxis treatment must be widely implemented and closely monitored during routine management of PLHIV irrespective of ART status. Additionally, AIDS-related ODs diagnostic capabilities require strengthening at all existing healthcare facilities initiating and monitoring antiretroviral therapy.

## Data Availability

All relevant data generated and analysed in the current study are available from the corresponding author, HHS, upon reasonable request.

## Authors’ contributions

HHS conceived the study, participated in the study design, data collection, analysis, and manuscript writing.

## Acknowledgements

The author is grateful to all gate keepers and research teams from all 8 research sites: Nkhoma Hospital, Lighthouse Trust & Martin Preus Centre, Partners-in-Hope, Mzuzu Hospital, DREAM Centre, Chiradzulu Hospital, QECH, and Thyolo Hospital ART facilities for their undivided support towards this study.

## Availability of data and materials

All relevant data generated or analysed during the current study are available from the corresponding author, HHS, on reasonable request.

## Ethical approval and consent to participate

The study obtained ethical approval from the University of Warwick Research Ethics Committee; The Biomedical and Scientific Research Ethics Committee (BSREC) [BSREC approval number: REGO-2015-1576], and from Malawi Research Ethics Committee; The National Health Sciences Research Committee (NHSRC) in the Ministry of Health [NHSRC approval number: NHSRC # 16/3/1555]. Approval from NHSRC was granted after obtaining “Letters of No Objection” from each of the 8 data collection centres. The need for consent to participate was waived off due to the nature and source of study’s data.

## Supplementary materials

None.

## APPENDIX TABLES

**Appendix A, Table 4:** Factors Associated with Manifestation of Bacterial and Fungal Opportunistic Diseases in Adults Living with HIV in Malawi; 2004-2015.

| Characteristics | Bacterial Conditions |  |  |  | Fungal Conditions |  |  |  |
| --- | --- | --- | --- | --- | --- | --- | --- | --- |
|  | Univariable findings |  | Multivariable findings |  | Univariable findings |  | Multivariable findings |  |
|  | OR (95% CI) | *p-value* | aOR (95% CI) | *p-value* | OR (95% CI) | *p-value* | aOR (95% CI) | *p-value* |
| Care type (*n*=9953) |  |  |  |  |  |  |  |  |
| Primary care | 1.00 (ref) |  |  |  |  |  |  |  |
| Secondary care | 1.38 (1.13, 1.69) | 0.002 | 1.34 (1.08, 1.66) | 0.008 | 1.09 (1.30, 3.91) | 0.007 | 1.33 (1.26, 4.91) | 0.016 |
| Tertiary care | 1.11 (0.92, 1.35) | 0.262 | 1.08 (0.88, 1.31) | 0.468 | 4.43 (1.41, 13.97) | 0.008 | 4.42 (1.38, 14.07) | 0.004 |
| Gender (*n*=9953) |  |  |  |  |  |  |  |  |
| Female | 1.00 (ref) |  |  |  |  |  |  |  |
| Male | 1.00 (0.91, 1.10) | 1.00 | 1.05 (0.93, 1.18) | 0.4 | 0.98 (0.89, 1.09) | 0.727 | 0.97 (0.85, 1.10) | 0.596 |
| Age-group (*n*=9950) |  |  |  |  |  |  |  |  |
| 15-19 years | 1.00 (ref) |  |  |  |  |  |  |  |
| 20-39 years | 1.17 (0.89, 1.52) | 0.255 | 1.26 (0.94, 1.70) | 0.121 | 1.16 (1.07, 1.55) | 0.031 | 1.07 (1.03, 1.47) | 0.017 |
| 40-54 years | 1.13 (0.86, 1.48) | 0.375 | 1.19 (0.88, 1.62) | 0.25 | 1.08 (1.01, 1.44) | 0.051 | 1.02 (1.01, 1.42) | 0.008 |
| 55+ years | 1.13 (0.85, 1.51) | 0.408 | 1.14 (0.83, 1.58) | 0.422 | 1.13 (0.83, 1.54) | 0.348 | 1.06 (0.75, 1.50) | 0.738 |
| Residential area (*n*=9953) |  |  |  |  |  |  |  |  |
| Rural | 1.00 (ref) |  |  |  |  |  |  |  |
| Semi-urban | 1.05 (0.90, 1.24) | 0.524 | 1.16 (0.97, 1.38) | 0.096 | 1.45 (1.38, 4.33) | 0.006 | 1.52 (1.48, 2.76) | 0.019 |
| Urban | 1.03 (0.91, 1.16) | 0.675 | 1.15 (1.01, 1.30) | 0.032 | 1.38 (1.24, 2.28) | 0.009 | 1.15 (1.09, 1.93) | 0.006 |
| Regional area (*n*=9953) |  |  |  |  |  |  |  |  |
| Central region | 1.00 (ref) |  |  |  |  |  |  |  |
| Northern Region | 0.72 (0.62, 0.85) | 0.0001 | 0.79 (0.67, 0.94) | 0.006 | 0.89 (0.75, 1.05) | 0.153 | 0.89 (0.75, 1.06) | 0.182 |
| Southern Region | 1.12 (1.00, 1.25) | 0.05 | 1.09 (1.07, 1.23) | 0.033 | 1.00 (0.89, 1.13) | 0.991 | 1.00 (0.89, 1.14) | 0.947 |
| Marital status (*n*=9949) |  |  |  |  |  |  |  |  |
| Previously married | 1.00 (ref) |  |  |  |  |  |  |  |
| Married (partnership) | 1.01 (0.88, 1.16) | 0.888 | 1.00 (0.87, 1.16) | 0.98 | 1.10 (1.05, 1.28) | 0.023 | 1.09 (1.03, 1.28) | 0.025 |
| Single (never married) | 1.27 (1.03, 1.57) | 0.023 | 1.51 (1.20, 1.90) | 0.001 | 1.00 (0.79, 1.27) | 0.971 | 1.04 (0.80, 1.36) | 0.751 |
| Education level (*n*=9953) |  |  |  |  |  |  |  |  |
| No education | 1.00 (ref) |  |  |  |  |  |  |  |
| Primary | 0.89 (0.74, 1.08) | 0.242 | 0.93 (0.76, 1.14) | 0.506 | 0.91 (0.74, 0.97) | 0.035 | 0.97 (0.79, 0.99) | 0.007 |
| Secondary | 0.88 (0.72, 1.06) | 0.176 | 0.91 (0.74, 1.12) | 0.354 | 0.86 (0.70, 0.91) | 0.013 | 0.93 (0.75, 0.96) | 0.053 |
| Higher | 0.96 (0.79, 1.17) | 0.682 | 1.01 (0.82, 1.25) | 0.923 | 0.95 (0.78, 0.97) | 0.006 | 1.04 (1.02, 1.30) | 0.037 |
| Employment status (*n*=9953) |  |  |  |  |  |  |  |  |
| Employed | 1.00 (ref) |  |  |  |  |  |  |  |
| Self-employed | 0.89 (0.81, 0.98) | 0.02 | 0.88 (0.79, 0.97) | 0.011 | 0.91 (0.82, 1.01) | 0.067 | 0.93 (0.84, 0.94) | 0.022 |
| Occupation type (*n*=9953) |  |  |  |  |  |  |  |  |
| Manual jobs | 1.00 (ref) |  |  |  |  |  |  |  |
| Other job types | 0.98 (0.86, 1.12) | 0.73 | 0.94 (0.81, 1.09) | 0.394 | 1.10 (0.95, 1.27) | 0.206 | 1.06 (0.91, 1.24) | 0.466 |
| White collar | 0.97 (0.85, 1.12) | 0.723 | 0.93 (0.79, 1.08) | 0.331 | 1.06 (0.91, 1.23) | 0.461 | 0.99 (0.84, 1.18) | 0.934 |
| ART initiation criteria (*n*=9953)<br>CD4 count | 1.00 (ref) |  |  |  |  |  |  |  |
| Option-B+/PMTCT | 0.99 (0.68, 1.42) | 0.942 | 1.03 (0.71, 1.49) | 0.87 | 1.11 (0.78, 1.57) | 0.574 | 1.11 (0.78, 1.58) | 0.567 |
| WHO III | 2.13 (1.87, 2.42) | 0.0001 | 2.22 (1.93, 2.55) | 0.0001 | 1.25 (1.10, 1.43) | 0.001 | 1.18 (1.03, 1.36) | 0.02 |
| WHO IV | 1.97 (1.70, 2.28) | 0.0001 | 1.98 (1.70, 2.31) | 0.0001 | 1.89 (1.63, 2.18) | 0.0001 | 1.85 (1.59, 2.16) | 0.0001 |
| CD4-initial results (*n*=9944)<br>0-250 cells/μL | 1.00 (ref) | | | | | | | |
| >250 cells/μL | 1.00 (0.91, 1.11) | 0.94 | 1.02 (0.91, 1.13) | 0.768 | 0.61 (0.54, 0.69) | 0.0001 | 0.59 (0.52, 0.66) | 0.0001 |
| CD4-follow-up results (*n*=9951)<br>100-500 cells/μL | 1.00 (ref) | | | | | | | |
| 501-1000 cells/μL | 1.00 (0.90, 1.11) | 0.968 | 1.04 (0.90, 1.20) | 0.612 | 0.65 (0.45, 0.95) | 0.033 | 0.61 (0.38, 0.98) | 0.025 |
| >1000 cells/μL | 0.88 (0.76, 0.92) | 0.051 | 0.90 (0.76, 0.98) | 0.027 | 0.75 (0.45, 1.24) | 0.082 | 0.68 (0.38, 0.73) | 0.009 |
| VL-initial results (*n*=9945)<br><3.0 Log<sub>10</sub> copies/mL | 1.00 (ref) |  |  |  |  |  |  |  |
| >3.0 Log<sub>10</sub> copies/mL | 1.66 (1.24, 2.22) | 0.001 | 1.92 (1.68, 2.27) | 0.023 | 1.70 (1.23, 2.35) | 0.001 | 1.25 (0.89, 1.76) | 0.196 |
| VL-follow-up results (*n*=9953)<br><1.6 Log<sub>10</sub> copies/mL |  |  |  |  |  |  |  |  |
| 1.6-3.0 Log<sub>10</sub> copies/mL | 0.97 (0.86, 1.08) | 0.543 | 0.97 (0.85, 1.10) | 0.59 | 1.15 (1.02, 1.30) | 0.025 | 1.14 (1.00, 1.31) | 0.052 |
| >3.0 Log<sub>10</sub> copies/mL | 0.96 (0.86, 1.08) | 0.489 | 0.87 (0.74, 1.01) | 0.073 | 1.12 (0.99, 1.26) | 0.076 | 1.22 (1.04, 1.44) | 0.016 |
| Current ART regimen (*n*=9953)<br>Nevirapine-based | 1.00 (ref) |  |  |  |  |  |  |  |
| Efavirenz-based | 1.10 (0.98, 1.23) | 0.091 | 1.08 (0.96, 1.21) | 0.206 | 1.08 (0.96, 1.22) | 0.202 | 1.08 (0.96, 1.22) | 0.215 |
| Boosted-Ritonavir | 1.41 (1.09, 1.83) | 0.01 | 1.39 (1.06, 1.81) | 0.016 | 0.80 (0.58, 1.10) | 0.174 | 0.78 (0.57, 1.09) | 0.142 |
| Non-std. | 1.21 (0.81, 1.83) | 0.354 | 1.26 (0.83, 1.91) | 0.277 | 0.68 (0.41, 1.15) | 0.151 | 0.69 (0.41, 1.17) | 0.167 |
| HIV duration (*n*=9953)<br>1-2 years | 1.00 (ref) |  |  |  |  |  |  |  |
| 3-5 years | 1.15 (0.99, 1.35) | 0.075 | 1.14 (0.90, 1.44) | 0.267 | 0.90 (0.76, 1.06) | 0.194 | 0.91 (0.71, 1.17) | 0.469 |
| 6-10 years | 1.01 (0.87, 1.17) | 0.918 | 1.02 (0.76, 1.38) | 0.88 | 0.90 (0.78, 1.05) | 0.196 | 0.95 (0.68, 1.32) | 0.751 |
| >10 years | 1.05 (0.88, 1.26) | 0.553 | 1.09 (0.72, 1.66) | 0.676 | 0.80 (0.66, 0.96) | 0.018 | 0.89 (0.56, 0.94) | 0.017 |
| ART duration (*n*=9953)<br>1-2 years | 1.00 (ref) |  |  |  |  |  |  |  |
| 3-5 years | 1.04 (0.91, 1.18) | 0.571 | 1.02 (0.83, 1.24) | 0.881 | 0.94 (0.82, 1.07) | 0.347 | 0.99 (0.79, 1.24) | 0.932 |
| 6-10 years | 0.90 (0.80, 1.02) | 0.106 | 0.97 (0.74, 1.27) | 0.819 | 0.92 (0.81, 1.05) | 0.238 | 0.98 (0.73, 1.32) | 0.899 |
| >10 years | 0.88 (0.71, 1.09) | 0.242 | 0.91 (0.62, 1.35) | 0.649 | 0.78 (0.61, 0.98) | 0.037 | 0.89 (0.58, 0.93) | 0.011 |
| Clinical latency (*n*=9953)<br>1-2 years | 1.00 (ref) |  |  |  |  |  |  |  |
| 3-5 years | 1.10 (0.99, 1.23) | 0.084 | 1.05 (0.90, 1.22) | 0.532 | 0.97 (0.85, 1.09) | 0.579 | 1.01 (0.86, 1.19) | 0.91 |
| > 5 years | 1.09 (0.91, 1.31) | 0.328 | 1.00 (0.75, 1.32) | 0.991 | 0.91 (0.75, 1.11) | 0.355 | 1.00 (0.73, 1.36) | 0.995 |
| Treatment outcomes (*n*=9953)<br>Treatment adherent | 1.00 (ref) |  |  |  |  |  |  |  |
| Defaulters & LTFU | 1.12 (1.00, 1.26) | 0.051 | 1.10 (1.07, 1.24) | 0.053 | 1.02 (0.90, 1.16) | 0.109 | 1.19 (1.14, 1.90) | 0.017 |
| Died | 1.29 (1.12, 1.48) | 0.001 | 1.24 (1.02, 1.49) | 0.031 | 0.91 (0.77, 1.07) | 0.258 | 1.78 (1.64, 1.97) | 0.022 |

**Appendix B, Table 5:**
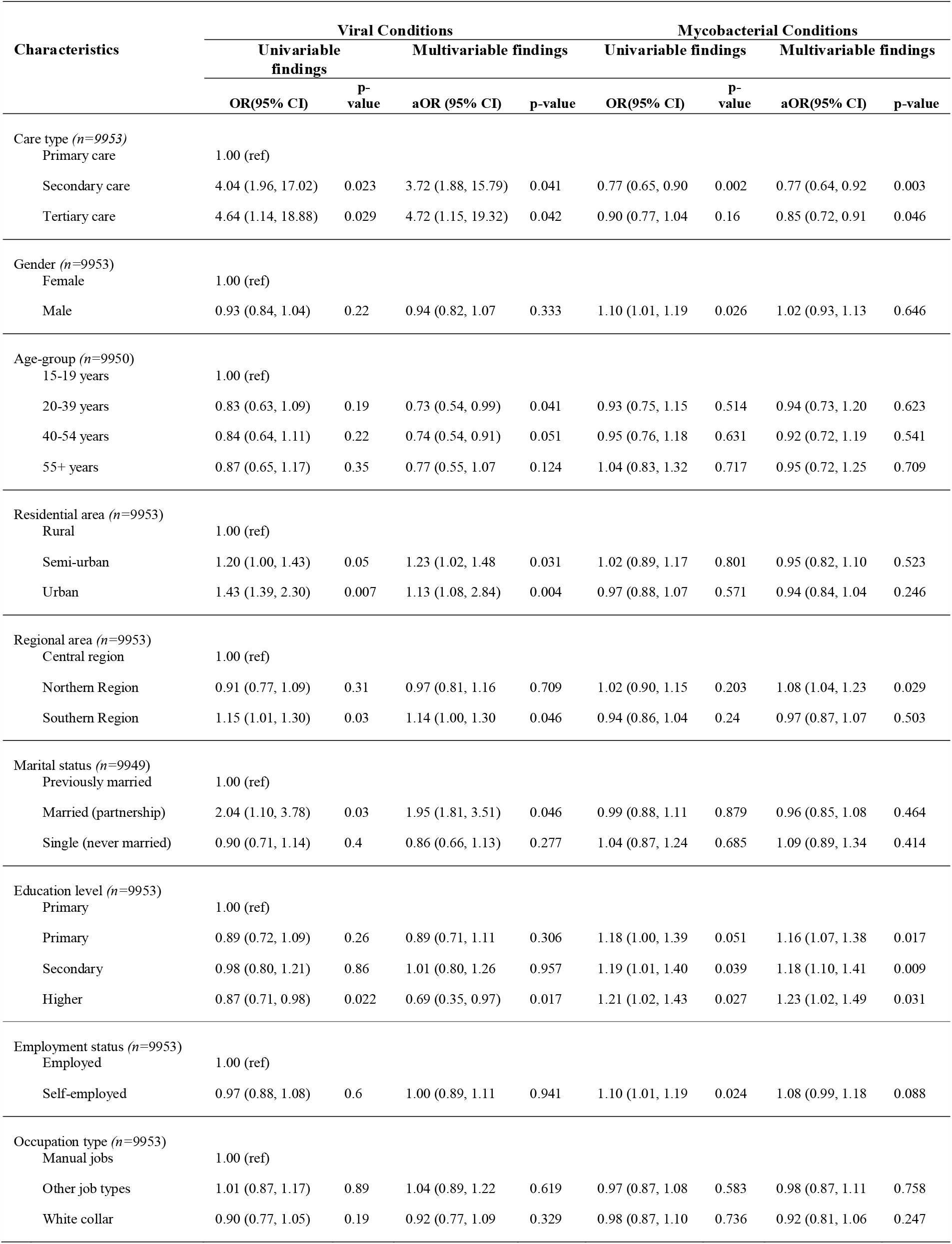
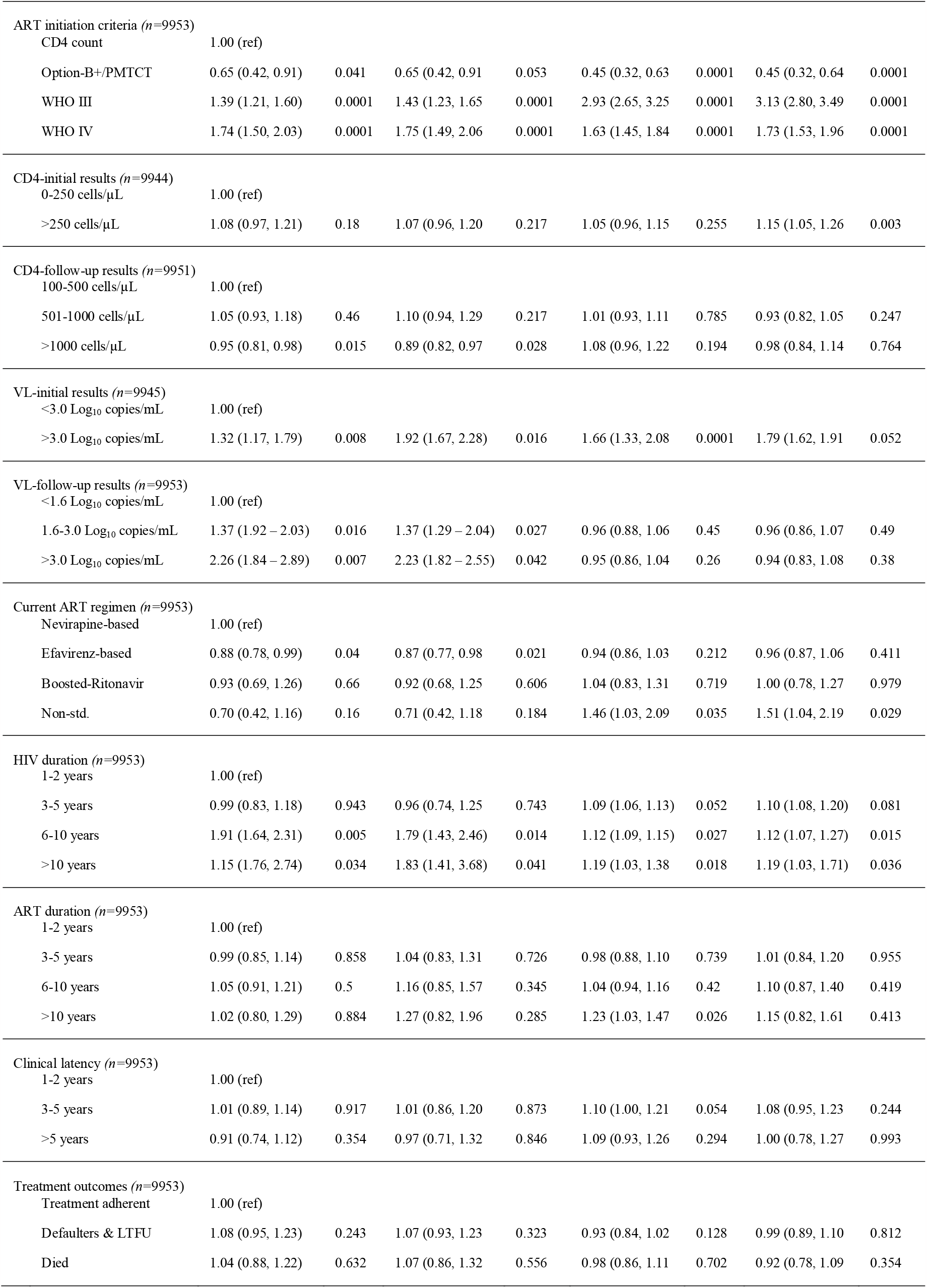
Factors Associated with Manifestation of Viral and Mycobacterial Opportunistic Diseases in Adults Living with HIV in Malawi; 2004-2015.

| Characteristics | Viral Conditions |  |  |  | Mycobacterial Conditions |  |  |  |
| --- | --- | --- | --- | --- | --- | --- | --- | --- |
|  | Univariable findings |  | Multivariable findings |  | Univariable findings |  | Multivariable findings |  |
|  | OR(95% CI) | p-value | aOR (95% CI) | p-value | OR(95% CI) | p-value | aOR(95% CI) | p-value |
| Care type (*n*=9953) |  |  |  |  |  |  |  |  |
| Primary care | 1.00 (ref) |  |  |  |  |  |  |  |
| Secondary care | 4.04 (1.96, 17.02) | 0.023 | 3.72 (1.88, 15.79) | 0.041 | 0.77 (0.65, 0.90) | 0.002 | 0.77 (0.64, 0.92) | 0.003 |
| Tertiary care | 4.64 (1.14, 18.88) | 0.029 | 4.72 (1.15, 19.32) | 0.042 | 0.90 (0.77, 1.04) | 0.16 | 0.85 (0.72, 0.91) | 0.046 |
| Gender (*n*=9953) |  |  |  |  |  |  |  |  |
| Female | 1.00 (ref) |  |  |  |  |  |  |  |
| Male | 0.93 (0.84, 1.04) | 0.22 | 0.94 (0.82, 1.07) | 0.333 | 1.10 (1.01, 1.19) | 0.026 | 1.02 (0.93, 1.13) | 0.646 |
| Age-group (*n*=9950) |  |  |  |  |  |  |  |  |
| 15-19 years | 1.00 (ref) |  |  |  |  |  |  |  |
| 20-39 years | 0.83 (0.63, 1.09) | 0.19 | 0.73 (0.54, 0.99) | 0.041 | 0.93 (0.75, 1.15) | 0.514 | 0.94 (0.73, 1.20) | 0.623 |
| 40-54 years | 0.84 (0.64, 1.11) | 0.22 | 0.74 (0.54, 0.91) | 0.051 | 0.95 (0.76, 1.18) | 0.631 | 0.92 (0.72, 1.19) | 0.541 |
| 55+ years | 0.87 (0.65, 1.17) | 0.35 | 0.77 (0.55, 1.07) | 0.124 | 1.04 (0.83, 1.32) | 0.717 | 0.95 (0.72, 1.25) | 0.709 |
| Residential area (*n*=9953) |  |  |  |  |  |  |  |  |
| Rural | 1.00 (ref) |  |  |  |  |  |  |  |
| Semi-urban | 1.20 (1.00, 1.43) | 0.05 | 1.23 (1.02, 1.48) | 0.031 | 1.02 (0.89, 1.17) | 0.801 | 0.95 (0.82, 1.10) | 0.523 |
| Urban | 1.43 (1.39, 2.30) | 0.007 | 1.13 (1.08, 2.84) | 0.004 | 0.97 (0.88, 1.07) | 0.571 | 0.94 (0.84, 1.04) | 0.246 |
| Regional area (*n*=9953) |  |  |  |  |  |  |  |  |
| Central region | 1.00 (ref) |  |  |  |  |  |  |  |
| Northern Region | 0.91 (0.77, 1.09) | 0.31 | 0.97 (0.81, 1.16) | 0.709 | 1.02 (0.90, 1.15) | 0.203 | 1.08 (1.04, 1.23) | 0.029 |
| Southern Region | 1.15 (1.01, 1.30) | 0.03 | 1.14 (1.00, 1.30) | 0.046 | 0.94 (0.86, 1.04) | 0.24 | 0.97 (0.87, 1.07) | 0.503 |
| Marital status (*n*=9949) |  |  |  |  |  |  |  |  |
| Previously married | 1.00 (ref) |  |  |  |  |  |  |  |
| Married (partnership) | 2.04 (1.10, 3.78) | 0.03 | 1.95 (1.81, 3.51) | 0.046 | 0.99 (0.88, 1.11) | 0.879 | 0.96 (0.85, 1.08) | 0.464 |
| Single (never married) | 0.90 (0.71, 1.14) | 0.4 | 0.86 (0.66, 1.13) | 0.277 | 1.04 (0.87, 1.24) | 0.685 | 1.09 (0.89, 1.34) | 0.414 |
| Education level (*n*=9953) |  |  |  |  |  |  |  |  |
| Primary | 1.00 (ref) |  |  |  |  |  |  |  |
| Primary | 0.89 (0.72, 1.09) | 0.26 | 0.89 (0.71, 1.11) | 0.306 | 1.18 (1.00, 1.39) | 0.051 | 1.16 (1.07, 1.38) | 0.017 |
| Secondary | 0.98 (0.80, 1.21) | 0.86 | 1.01 (0.80, 1.26) | 0.957 | 1.19 (1.01, 1.40) | 0.039 | 1.18 (1.10, 1.41) | 0.009 |
| Higher | 0.87 (0.71, 0.98) | 0.022 | 0.69 (0.35, 0.97) | 0.017 | 1.21 (1.02, 1.43) | 0.027 | 1.23 (1.02, 1.49) | 0.031 |
| Employment status (*n*=9953) |  |  |  |  |  |  |  |  |
| Employed | 1.00 (ref) |  |  |  |  |  |  |  |
| Self-employed | 0.97 (0.88, 1.08) | 0.6 | 1.00 (0.89, 1.11) | 0.941 | 1.10 (1.01, 1.19) | 0.024 | 1.08 (0.99, 1.18) | 0.088 |
| Occupation type (*n*=9953) |  |  |  |  |  |  |  |  |
| Manual jobs | 1.00 (ref) |  |  |  |  |  |  |  |
| Other job types | 1.01 (0.87, 1.17) | 0.89 | 1.04 (0.89, 1.22) | 0.619 | 0.97 (0.87, 1.08) | 0.583 | 0.98 (0.87, 1.11) | 0.758 |
| White collar | 0.90 (0.77, 1.05) | 0.19 | 0.92 (0.77, 1.09) | 0.329 | 0.98 (0.87, 1.10) | 0.736 | 0.92 (0.81, 1.06) | 0.247 |
| ART initiation criteria (*n*=9953) |  |  |  |  |  |  |  |  |
| CD4 count | 1.00 (ref) |  |  |  |  |  |  |  |
| Option-B+/PMTCT | 0.65 (0.42, 0.91) | 0.041 | 0.65 (0.42, 0.91) | 0.053 | 0.45 (0.32, 0.63) | 0.0001 | 0.45 (0.32, 0.64) | 0.0001 |
| WHO III | 1.39 (1.21, 1.60) | 0.0001 | 1.43 (1.23, 1.65) | 0.0001 | 2.93 (2.65, 3.25) | 0.0001 | 3.13 (2.80, 3.49) | 0.0001 |
| WHO IV | 1.74 (1.50, 2.03) | 0.0001 | 1.75 (1.49, 2.06) | 0.0001 | 1.63 (1.45, 1.84) | 0.0001 | 1.73 (1.53, 1.96) | 0.0001 |
| CD4-initial results (*n*=9944) |  |  |  |  |  |  |  |  |
| 0-250 cells/μL | 1.00 (ref) |  |  |  |  |  |  |  |
| >250 cells/μL | 1.08 (0.97, 1.21) | 0.18 | 1.07 (0.96, 1.20) | 0.217 | 1.05 (0.96, 1.15) | 0.255 | 1.15 (1.05, 1.26) | 0.003 |
| CD4-follow-up results (*n*=9951) |  |  |  |  |  |  |  |  |
| 100-500 cells/μL | 1.00 (ref) |  |  |  |  |  |  |  |
| 501-1000 cells/μL | 1.05 (0.93, 1.18) | 0.46 | 1.10 (0.94, 1.29) | 0.217 | 1.01 (0.93, 1.11) | 0.785 | 0.93 (0.82, 1.05) | 0.247 |
| >1000 cells/μL | 0.95 (0.81, 0.98) | 0.015 | 0.89 (0.82, 0.97) | 0.028 | 1.08 (0.96, 1.22) | 0.194 | 0.98 (0.84, 1.14) | 0.764 |
| VL-initial results (*n*=9945) |  |  |  |  |  |  |  |  |
| <3.0 Log<sub>10</sub> copies/mL | 1.00 (ref) |  |  |  |  |  |  |  |
| >3.0 Log<sub>10</sub> copies/mL | 1.32 (1.17, 1.79) | 0.008 | 1.92 (1.67, 2.28) | 0.016 | 1.66 (1.33, 2.08) | 0.0001 | 1.79 (1.62, 1.91) | 0.052 |
| VL-follow-up results (*n*=9953) |  |  |  |  |  |  |  |  |
| <1.6 Log<sub>10</sub> copies/mL | 1.00 (ref) |  |  |  |  |  |  |  |
| 1.6-3.0 Log<sub>10</sub> copies/mL | 1.37 (1.92 – 2.03) | 0.016 | 1.37 (1.29 – 2.04) | 0.027 | 0.96 (0.88, 1.06) | 0.45 | 0.96 (0.86, 1.07) | 0.49 |
| >3.0 Log<sub>10</sub> copies/mL | 2.26 (1.84 – 2.89) | 0.007 | 2.23 (1.82 – 2.55) | 0.042 | 0.95 (0.86, 1.04) | 0.26 | 0.94 (0.83, 1.08) | 0.38 |
| Current ART regimen (*n*=9953) |  |  |  |  |  |  |  |  |
| Nevirapine-based | 1.00 (ref) |  |  |  |  |  |  |  |
| Efavirenz-based | 0.88 (0.78, 0.99) | 0.04 | 0.87 (0.77, 0.98) | 0.021 | 0.94 (0.86, 1.03) | 0.212 | 0.96 (0.87, 1.06) | 0.411 |
| Boosted-Ritonavir | 0.93 (0.69, 1.26) | 0.66 | 0.92 (0.68, 1.25) | 0.606 | 1.04 (0.83, 1.31) | 0.719 | 1.00 (0.78, 1.27) | 0.979 |
| Non-std. | 0.70 (0.42, 1.16) | 0.16 | 0.71 (0.42, 1.18) | 0.184 | 1.46 (1.03, 2.09) | 0.035 | 1.51 (1.04, 2.19) | 0.029 |
| HIV duration (*n*=9953) |  |  |  |  |  |  |  |  |
| 1-2 years | 1.00 (ref) |  |  |  |  |  |  |  |
| 3-5 years | 0.99 (0.83, 1.18) | 0.943 | 0.96 (0.74, 1.25) | 0.743 | 1.09 (1.06, 1.13) | 0.052 | 1.10 (1.08, 1.20) | 0.081 |
| 6-10 years | 1.91 (1.64, 2.31) | 0.005 | 1.79 (1.43, 2.46) | 0.014 | 1.12 (1.09, 1.15) | 0.027 | 1.12 (1.07, 1.27) | 0.015 |
| >10 years | 1.15 (1.76, 2.74) | 0.034 | 1.83 (1.41, 3.68) | 0.041 | 1.19 (1.03, 1.38) | 0.018 | 1.19 (1.03, 1.71) | 0.036 |
| ART duration (*n*=9953) |  |  |  |  |  |  |  |  |
| 1-2 years | 1.00 (ref) |  |  |  |  |  |  |  |
| 3-5 years | 0.99 (0.85, 1.14) | 0.858 | 1.04 (0.83, 1.31) | 0.726 | 0.98 (0.88, 1.10) | 0.739 | 1.01 (0.84, 1.20) | 0.955 |
| 6-10 years | 1.05 (0.91, 1.21) | 0.5 | 1.16 (0.85, 1.57) | 0.345 | 1.04 (0.94, 1.16) | 0.42 | 1.10 (0.87, 1.40) | 0.419 |
| >10 years | 1.02 (0.80, 1.29) | 0.884 | 1.27 (0.82, 1.96) | 0.285 | 1.23 (1.03, 1.47) | 0.026 | 1.15 (0.82, 1.61) | 0.413 |
| Clinical latency (*n*=9953) |  |  |  |  |  |  |  |  |
| 1-2 years | 1.00 (ref) |  |  |  |  |  |  |  |
| 3-5 years | 1.01 (0.89, 1.14) | 0.917 | 1.01 (0.86, 1.20) | 0.873 | 1.10 (1.00, 1.21) | 0.054 | 1.08 (0.95, 1.23) | 0.244 |
| >5 years | 0.91 (0.74, 1.12) | 0.354 | 0.97 (0.71, 1.32) | 0.846 | 1.09 (0.93, 1.26) | 0.294 | 1.00 (0.78, 1.27) | 0.993 |
| Treatment outcomes (*n*=9953) |  |  |  |  |  |  |  |  |
| Treatment adherent | 1.00 (ref) |  |  |  |  |  |  |  |
| Defaulters & LTFU | 1.08 (0.95, 1.23) | 0.243 | 1.07 (0.93, 1.23) | 0.323 | 0.93 (0.84, 1.02) | 0.128 | 0.99 (0.89, 1.10) | 0.812 |
| Died | 1.04 (0.88, 1.22) | 0.632 | 1.07 (0.86, 1.32) | 0.556 | 0.98 (0.86, 1.11) | 0.702 | 0.92 (0.78, 1.09) | 0.354 |

**Appendix C, Table 6:**
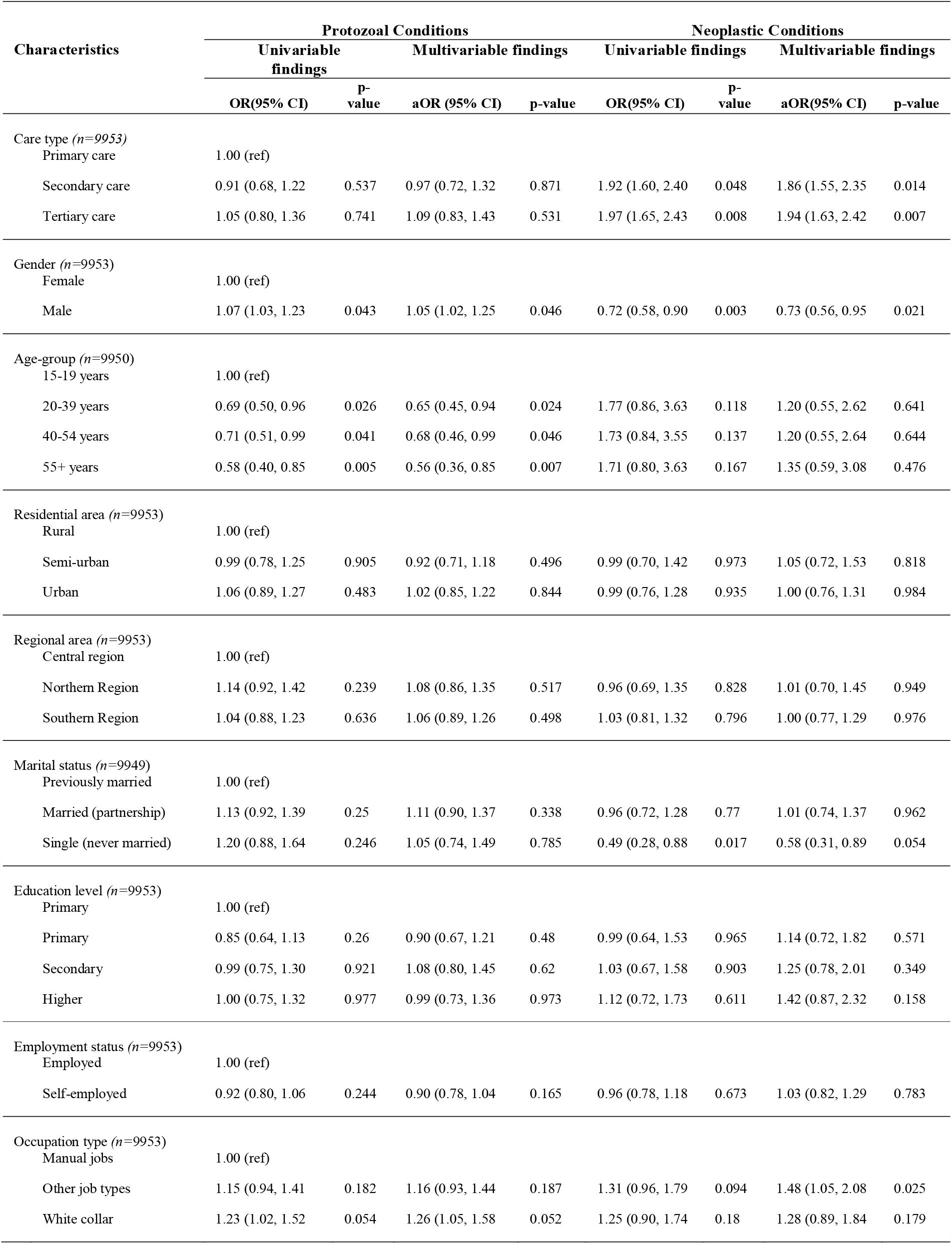
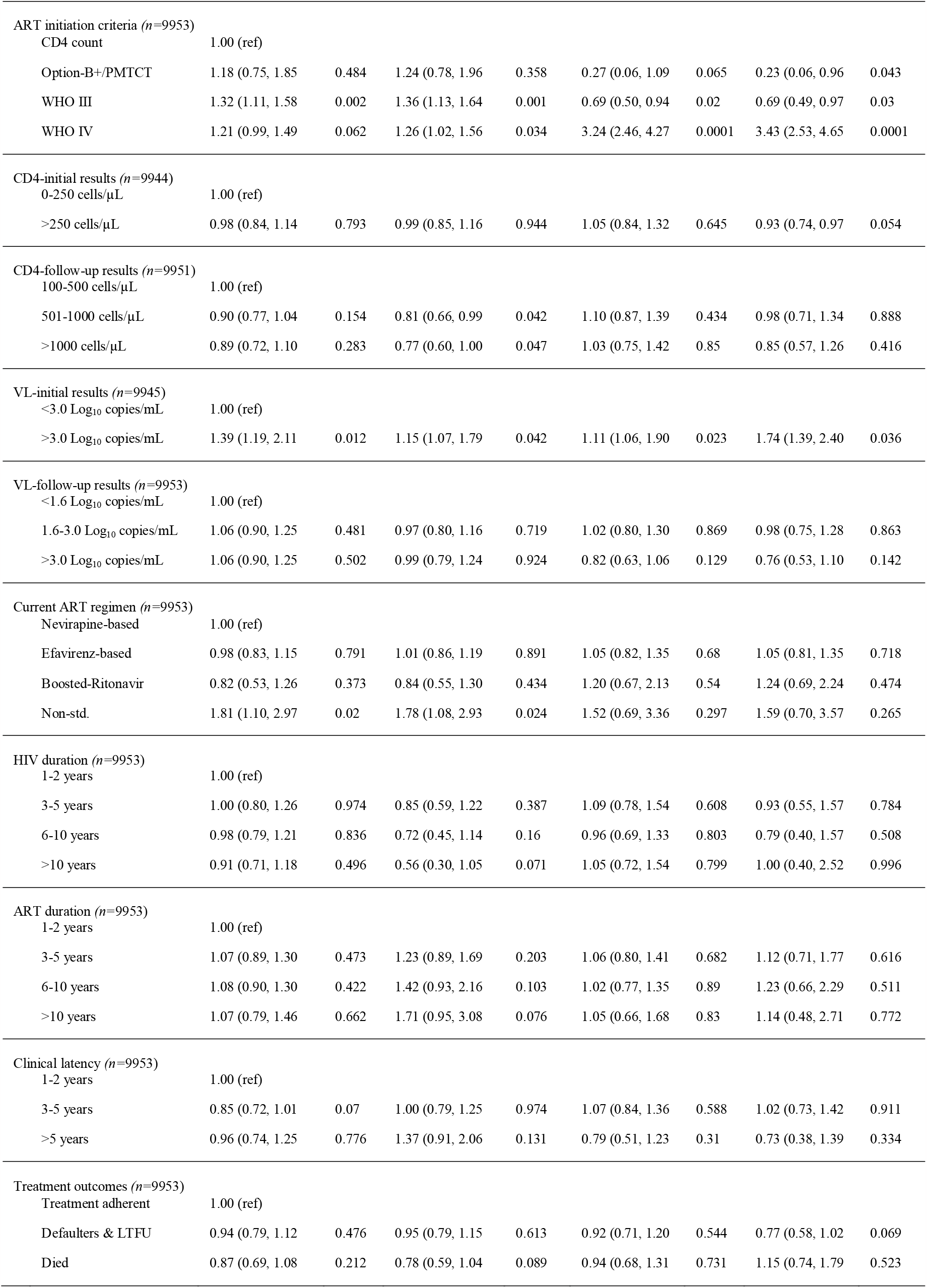
Factors Associated with Manifestation of Protozoal (Parasitic) and Neoplastic Opportunistic Diseases in Adults Living with HIV in Malawi; 2004-2015.

| Characteristics | Protozoal Conditions |  |  |  | Neoplastic Conditions |  |  |  |
| --- | --- | --- | --- | --- | --- | --- | --- | --- |
|  | Univariable findings |  | Multivariable findings |  | Univariable findings |  | Multivariable findings |  |
|  | OR(95% CI) | p-value | aOR (95% CI) | p-value | OR(95% CI) | p-value | aOR(95% CI) | p-value |
| Care type (*n*=9953) |  |  |  |  |  |  |  |  |
| Primary care | 1.00 (ref) |  |  |  |  |  |  |  |
| Secondary care | 0.91 (0.68, 1.22) | 0.537 | 0.97 (0.72, 1.32) | 0.871 | 1.92 (1.60, 2.40) | 0.048 | 1.86 (1.55, 2.35) | 0.014 |
| Tertiary care | 1.05 (0.80, 1.36) | 0.741 | 1.09 (0.83, 1.43) | 0.531 | 1.97 (1.65, 2.43) | 0.008 | 1.94 (1.63, 2.42) | 0.007 |
| Gender (*n*=9953) |  |  |  |  |  |  |  |  |
| Female | 1.00 (ref) |  |  |  |  |  |  |  |
| Male | 1.07 (1.03, 1.23) | 0.043 | 1.05 (1.02, 1.25) | 0.046 | 0.72 (0.58, 0.90) | 0.003 | 0.73 (0.56, 0.95) | 0.021 |
| Age-group (*n*=9950) |  |  |  |  |  |  |  |  |
| 15-19 years | 1.00 (ref) |  |  |  |  |  |  |  |
| 20-39 years | 0.69 (0.50, 0.96) | 0.026 | 0.65 (0.45, 0.94) | 0.024 | 1.77 (0.86, 3.63) | 0.118 | 1.20 (0.55, 2.62) | 0.641 |
| 40-54 years | 0.71 (0.51, 0.99) | 0.041 | 0.68 (0.46, 0.99) | 0.046 | 1.73 (0.84, 3.55) | 0.137 | 1.20 (0.55, 2.64) | 0.644 |
| 55+ years | 0.58 (0.40, 0.85) | 0.005 | 0.56 (0.36, 0.85) | 0.007 | 1.71 (0.80, 3.63) | 0.167 | 1.35 (0.59, 3.08) | 0.476 |
| Residential area (*n*=9953) |  |  |  |  |  |  |  |  |
| Rural | 1.00 (ref) |  |  |  |  |  |  |  |
| Semi-urban | 0.99 (0.78, 1.25) | 0.905 | 0.92 (0.71, 1.18) | 0.496 | 0.99 (0.70, 1.42) | 0.973 | 1.05 (0.72, 1.53) | 0.818 |
| Urban | 1.06 (0.89, 1.27) | 0.483 | 1.02 (0.85, 1.22) | 0.844 | 0.99 (0.76, 1.28) | 0.935 | 1.00 (0.76, 1.31) | 0.984 |
| Regional area (*n*=9953) |  |  |  |  |  |  |  |  |
| Central region | 1.00 (ref) |  |  |  |  |  |  |  |
| Northern Region | 1.14 (0.92, 1.42) | 0.239 | 1.08 (0.86, 1.35) | 0.517 | 0.96 (0.69, 1.35) | 0.828 | 1.01 (0.70, 1.45) | 0.949 |
| Southern Region | 1.04 (0.88, 1.23) | 0.636 | 1.06 (0.89, 1.26) | 0.498 | 1.03 (0.81, 1.32) | 0.796 | 1.00 (0.77, 1.29) | 0.976 |
| Marital status (*n*=9949) |  |  |  |  |  |  |  |  |
| Previously married | 1.00 (ref) |  |  |  |  |  |  |  |
| Married (partnership) | 1.13 (0.92, 1.39) | 0.25 | 1.11 (0.90, 1.37) | 0.338 | 0.96 (0.72, 1.28) | 0.77 | 1.01 (0.74, 1.37) | 0.962 |
| Single (never married) | 1.20 (0.88, 1.64) | 0.246 | 1.05 (0.74, 1.49) | 0.785 | 0.49 (0.28, 0.88) | 0.017 | 0.58 (0.31, 0.89) | 0.054 |
| Education level (*n*=9953) |  |  |  |  |  |  |  |  |
| Primary | 1.00 (ref) |  |  |  |  |  |  |  |
| Primary | 0.85 (0.64, 1.13) | 0.26 | 0.90 (0.67, 1.21) | 0.48 | 0.99 (0.64, 1.53) | 0.965 | 1.14 (0.72, 1.82) | 0.571 |
| Secondary | 0.99 (0.75, 1.30) | 0.921 | 1.08 (0.80, 1.45) | 0.62 | 1.03 (0.67, 1.58) | 0.903 | 1.25 (0.78, 2.01) | 0.349 |
| Higher | 1.00 (0.75, 1.32) | 0.977 | 0.99 (0.73, 1.36) | 0.973 | 1.12 (0.72, 1.73) | 0.611 | 1.42 (0.87, 2.32) | 0.158 |
| Employment status (*n*=9953) |  |  |  |  |  |  |  |  |
| Employed | 1.00 (ref) |  |  |  |  |  |  |  |
| Self-employed | 0.92 (0.80, 1.06) | 0.244 | 0.90 (0.78, 1.04) | 0.165 | 0.96 (0.78, 1.18) | 0.673 | 1.03 (0.82, 1.29) | 0.783 |
| Occupation type (*n*=9953) |  |  |  |  |  |  |  |  |
| Manual jobs | 1.00 (ref) |  |  |  |  |  |  |  |
| Other job types | 1.15 (0.94, 1.41) | 0.182 | 1.16 (0.93, 1.44) | 0.187 | 1.31 (0.96, 1.79) | 0.094 | 1.48 (1.05, 2.08) | 0.025 |
| White collar | 1.23 (1.02, 1.52) | 0.054 | 1.26 (1.05, 1.58) | 0.052 | 1.25 (0.90, 1.74) | 0.18 | 1.28 (0.89, 1.84) | 0.179 |

|  |  |  |  |  |  |  |  |  |  |
| --- | --- | --- | --- | --- | --- | --- | --- | --- | --- |
| ART initiation criteria (*n*=9953) |  |  |  |  |  |  |  |  |  |
| CD4 count | 1.00 (ref) |  |  |  |  |  |  |  |  |
| Option-B+/PMTCT | 1.18 (0.75, 1.85) | 0.484 | 1.24 (0.78, 1.96) | 0.358 | 0.27 (0.06, 1.09) | 0.065 | 0.23 (0.06, 0.96) | 0.043 |  |
| WHO III | 1.32 (1.11, 1.58) | 0.002 | 1.36 (1.13, 1.64) | 0.001 | 0.69 (0.50, 0.94) | 0.02 | 0.69 (0.49, 0.97) | 0.03 |  |
| WHO IV | 1.21 (0.99, 1.49) | 0.062 | 1.26 (1.02, 1.56) | 0.034 | 3.24 (2.46, 4.27) | 0.0001 | 3.43 (2.53, 4.65) | 0.0001 |  |
| CD4-initial results (*n*=9944) |  |  |  |  |  |  |  |  |  |
| 0-250 cells/μL | 1.00 (ref) | | | | | | | | |
| >250 cells/μL | 0.98 (0.84, 1.14) | 0.793 | 0.99 (0.85, 1.16) | 0.944 | 1.05 (0.84, 1.32) | 0.645 | 0.93 (0.74, 0.97) | 0.054 | |
| CD4-follow-up results (*n*=9951) |  |  |  |  |  |  |  |  |  |
| 100-500 cells/μL | 1.00 (ref) | | | | | | | | |
| 501-1000 cells/μL | 0.90 (0.77, 1.04) | 0.154 | 0.81 (0.66, 0.99) | 0.042 | 1.10 (0.87, 1.39) | 0.434 | 0.98 (0.71, 1.34) | 0.888 | |
| >1000 cells/μL | 0.89 (0.72, 1.10) | 0.283 | 0.77 (0.60, 1.00) | 0.047 | 1.03 (0.75, 1.42) | 0.85 | 0.85 (0.57, 1.26) | 0.416 | |
| VL-initial results (*n*=9945) |  |  |  |  |  |  |  |  |  |
| <3.0 Log<sub>10</sub> copies/mL | 1.00 (ref) |  |  |  |  |  |  |  |  |
| >3.0 Log<sub>10</sub> copies/mL | 1.39 (1.19, 2.11) | 0.012 | 1.15 (1.07, 1.79) | 0.042 | 1.11 (1.06, 1.90) | 0.023 | 1.74 (1.39, 2.40) | 0.036 |  |
| VL-follow-up results (*n*=9953) |  |  |  |  |  |  |  |  |  |
| <1.6 Log<sub>10</sub> copies/mL | 1.00 (ref) |  |  |  |  |  |  |  |  |
| 1.6-3.0 Log<sub>10</sub> copies/mL | 1.06 (0.90, 1.25) | 0.481 | 0.97 (0.80, 1.16) | 0.719 | 1.02 (0.80, 1.30) | 0.869 | 0.98 (0.75, 1.28) | 0.863 |  |
| >3.0 Log<sub>10</sub> copies/mL | 1.06 (0.90, 1.25) | 0.502 | 0.99 (0.79, 1.24) | 0.924 | 0.82 (0.63, 1.06) | 0.129 | 0.76 (0.53, 1.10) | 0.142 |  |
| Current ART regimen (*n*=9953) |  |  |  |  |  |  |  |  |  |
| Nevirapine-based | 1.00 (ref) |  |  |  |  |  |  |  |  |
| Efavirenz-based | 0.98 (0.83, 1.15) | 0.791 | 1.01 (0.86, 1.19) | 0.891 | 1.05 (0.82, 1.35) | 0.68 | 1.05 (0.81, 1.35) | 0.718 |  |
| Boosted-Ritonavir | 0.82 (0.53, 1.26) | 0.373 | 0.84 (0.55, 1.30) | 0.434 | 1.20 (0.67, 2.13) | 0.54 | 1.24 (0.69, 2.24) | 0.474 |  |
| Non-std. | 1.81 (1.10, 2.97) | 0.02 | 1.78 (1.08, 2.93) | 0.024 | 1.52 (0.69, 3.36) | 0.297 | 1.59 (0.70, 3.57) | 0.265 |  |
| HIV duration (*n*=9953) |  |  |  |  |  |  |  |  |  |
| 1-2 years | 1.00 (ref) |  |  |  |  |  |  |  |  |
| 3-5 years | 1.00 (0.80, 1.26) | 0.974 | 0.85 (0.59, 1.22) | 0.387 | 1.09 (0.78, 1.54) | 0.608 | 0.93 (0.55, 1.57) | 0.784 |  |
| 6-10 years | 0.98 (0.79, 1.21) | 0.836 | 0.72 (0.45, 1.14) | 0.16 | 0.96 (0.69, 1.33) | 0.803 | 0.79 (0.40, 1.57) | 0.508 |  |
| >10 years | 0.91 (0.71, 1.18) | 0.496 | 0.56 (0.30, 1.05) | 0.071 | 1.05 (0.72, 1.54) | 0.799 | 1.00 (0.40, 2.52) | 0.996 |  |
| ART duration (*n*=9953) |  |  |  |  |  |  |  |  |  |
| 1-2 years | 1.00 (ref) |  |  |  |  |  |  |  |  |
| 3-5 years | 1.07 (0.89, 1.30) | 0.473 | 1.23 (0.89, 1.69) | 0.203 | 1.06 (0.80, 1.41) | 0.682 | 1.12 (0.71, 1.77) | 0.616 |  |
| 6-10 years | 1.08 (0.90, 1.30) | 0.422 | 1.42 (0.93, 2.16) | 0.103 | 1.02 (0.77, 1.35) | 0.89 | 1.23 (0.66, 2.29) | 0.511 |  |
| >10 years | 1.07 (0.79, 1.46) | 0.662 | 1.71 (0.95, 3.08) | 0.076 | 1.05 (0.66, 1.68) | 0.83 | 1.14 (0.48, 2.71) | 0.772 |  |
| Clinical latency (*n*=9953) |  |  |  |  |  |  |  |  |  |
| 1-2 years | 1.00 (ref) |  |  |  |  |  |  |  |  |
| 3-5 years | 0.85 (0.72, 1.01) | 0.07 | 1.00 (0.79, 1.25) | 0.974 | 1.07 (0.84, 1.36) | 0.588 | 1.02 (0.73, 1.42) | 0.911 |  |
| >5 years | 0.96 (0.74, 1.25) | 0.776 | 1.37 (0.91, 2.06) | 0.131 | 0.79 (0.51, 1.23) | 0.31 | 0.73 (0.38, 1.39) | 0.334 |  |
| Treatment outcomes (*n*=9953) |  |  |  |  |  |  |  |  |  |
| Treatment adherent | 1.00 (ref) |  |  |  |  |  |  |  |  |
| Defaulters & LTFU | 0.94 (0.79, 1.12) | 0.476 | 0.95 (0.79, 1.15) | 0.613 | 0.92 (0.71, 1.20) | 0.544 | 0.77 (0.58, 1.02) | 0.069 |  |
| Died | 0.87 (0.69, 1.08) | 0.212 | 0.78 (0.59, 1.04) | 0.089 | 0.94 (0.68, 1.31) | 0.731 | 1.15 (0.74, 1.79) | 0.523 |  |

